# Prevalence and expressivity of loss of function mutations in the Melanocortin 4 Receptor (*MC4R*) in a UK birth cohort

**DOI:** 10.1101/2020.10.30.20220467

**Authors:** KH Wade, BYH Lam, A Melvin, W Pan, LJ Corbin, DA Hughes, K Rainbow, JH Chen, K Duckett, X Liu, J Mokrosiński, A Mörseburg, S Neaves, A Williamson, C Zhang, IS Farooqi, GSH Yeo, NJ Timpson, S O’Rahilly

**Author notes:** Corresponding authors **Corresponding authors:** Nicholas J Timpson, MRC Integrative Epidemiology Unit, University of Bristol, Oakfield House, Oakfield Grove, Bristol, BS8 2BN, UK, Stephen O’Rahilly, MRC Metabolic Diseases Unit, Wellcome Trust-MRC Institute of Metabolic Science, University of Cambridge, Cambridge, CB2 0QQ, UK. Equal contributions (KH Wade, BYH Lam, A Melvin, GSH Yeo, NJ Timpson, S O’Rahilly).

## Abstract

Mutations in the melanocortin 4 receptor gene (*MC4R*) have frequently been reported in severe early-onset human obesity but the prevalence and extent of phenotypic impact of such mutations are unclear. In a large UK birth cohort, we found that 17 of 5724 unrelated participants (∼1/337; 0.30%) were heterozygous for functionally deleterious mutations. At age 18 years, the mean difference in body weight, body mass index (BMI) and fat mass was 17.76kg (95% CI: 9.41, 26.10), 4.84kg/m^2^ (95% CI: 2.19, 7.49) and 14.78kg (95% CI: 8.56, 20.99), respectively, in carriers of loss of function (LoF) mutations compared to non-LoF carriers. Carriage of LoF mutations increased adiposity from as early as 5 years. *MC4R* LoF was associated with an impact on BMI at age 18 years that was approximately double that of a genome-wide polygenic risk score (comparing the upper 10^th^ and lower 90^th^ percentile). An extrapolation of incidence for *MC4R* LoF mutations from this birth cohort suggests that up to ∼200,000 people in the UK are likely to carry such mutations. This frequency, combined with the substantial impact of these variants on adiposity, has implications for public health, and drug development.

## INTRODUCTION

Mutations disrupting the leptin-melanocortin system have frequently been reported in severe, early-onset human obesity but the prevalence and extent of phenotypic impact of such mutations are unclear. The critical role of the leptin-melanocortin system in the long-term sensing of body fat stores was first established in the 1990s, with defects in this system resulting in obesity in rodents and humans^1-5^. Specifically, the melanocortin 4 receptor (*MC4R*) is a G-protein coupled, seven-transmembrane receptor expressed widely in the central nervous system^6,7^. The binding of its natural agonists, the pro-opiomelanocortin-derived melanocortin peptides, alpha and beta melanocyte-stimulating hormone (MSH), results in the suppression of food intake and the activation of a subset of autonomic neurons of the sympathetic nervous system^8-10^. Leptin acts on hypothalamic neurons to promote the release of melanocortins and suppress the secretion of the melanocortin antagonist, agouti-related peptide (AGRP)^9,11^.

Severe early-onset human obesity associated with mutations in genes encoding leptin or the leptin receptor are very rare and observed only in individuals homozygous for loss of function (LoF) mutations in those genes^3,5^. However, in the case of mutations in the *MC4R* gene, severe early-onset obesity has been reported in multiple affected members of several families who only carried heterozygote LoF mutations^4,12^. Subsequent studies reported more severe obesity in homozygotes, suggesting a semi-dominant form of inheritance^13^.

Many *MC4R* LoF mutations have been described in obese individuals and families^14,15^ and the severity of disruption of *MC4R* signalling resulting from such mutations has been reported to correlate with adiposity and degree of hyperphagia^16,17^. While early reports based on clinically ascertained cohorts suggested a high penetrance of early-onset obesity, subsequent studies of less highly selected patients demonstrated that the carriage of LoF mutations was not always associated with obesity^15,18^. For example, in a population-based cohort from Germany, Hinney et al., using a mutational scanning technique, reported a prevalence of LoF mutations in *MC4R* of ∼0.1%^15^. Stutzmann et al. reported a prevalence of *MC4R* LoF mutations of 1.7% in obese European adults and that obesity in carriers of the same mutation differed across generations within the same families, providing evidence for gene-environment interaction^18^. In a study of participants in UK Biobank based on high density single nucleotide polymorphism (SNP) genotyping, Turcot et al. reported that, while carriers of a rare (0.01%) non-sense mutations were ∼7kg heavier, on average, than non-carriers (for an average 1.7m tall individual), the majority of carriers of this mutation were not obese^19^.

Pharmacological agonists of *MC4R* are in clinical development for the therapy of obesity^20^. In a Phase 1b trial of one such drug, setmelanotide, obese participants wild-type (WT) at *MC4R* showed drug-induced weight loss, with those carrying homozygous LoF mutations being unresponsive and heterozygous carriers of a subset of *MC4R* mutations having an intermediate response^17^. As preventive efforts for metabolic disease are increasingly focusing on tackling obesity in childhood^21^, knowledge regarding the prevalence of *MC4R* LoF mutations and their impact on body composition and growth during the first decades of life will be increasingly important and relevant to future drug development.

In order to determine the prevalence of functionally impaired *MC4R* mutations and their clinical and phenotypic consequences throughout childhood, adolescence and early adult life in an unselected UK population, we determined the sequence of the *MC4R* gene in participants from the Avon Longitudinal Study of Parents and Children (ALSPAC), a birth cohort recruited in Bristol (UK) in 1990-92 and repeatedly followed up until early adulthood^22,23^. We characterised the signalling properties of all non-synonymous mutations that were found and studied their associations with relevant obesity-related traits.

## RESULTS

### Detection of MC4R mutations by pooled sequencing

ALSPAC is a birth cohort originally comprised of >75% of all pregnancies delivered in the Greater Bristol area from 1990-92. Whilst a specific cohort, ALSPAC represents a population-based sample with deep longitudinal phenotyping suitable for the dissection of *MC4R* mutation associations. Characteristics of the sequenced set of individuals and the complete ALSPAC cohort were similar (**Supplementary Table 1**), suggesting that the sequenced set were at least representative of the wider cohort, which is well described in terms of both demographic profile^22^ and attrition^24^.

**Table 1.** Sanger sequencing validated non-synonymous *MC4R* mutations identified in those sequenced in ALSPAC

| ***MC4R* variant** | **Genomic position (bp)** | **Codon change** | **Database accession(s)** | **Domain** | **No of carriers<sup>1</sup>** | **ALSPAC MAF** |
| --- | --- | --- | --- | --- | --- | --- |
| p.T5N<sup>2</sup> | Chr18:60372336 | aCc/aAc | rs752432398 | N-term | 1 | 0.0087% |
| p.T11A | Chr18:60372319 | Act/Gct | rs372794914 | N-term | 2 | 0.0175% |
| p.S30F | Chr18:60372261 | tCc/tTc | rs13447323 | N-term | 3 | 0.0262% |
| p.S36T | Chr18:60372244 | Tct/Act | rs954123325 | N-term | 1 | 0.0087% |
| p.T53I | Chr18:60372192 | aCt/aTt | rs141148170 | TM1 | 3 | 0.0262% |
| p.Y80C | Chr18:60372111 | tAc/tGc | rs1368643838 | TM2 | 1 | 0.0087% |
| p.S85I<sup>2</sup> | Chr18:60372096 | aGc/aTc | rs1420993856 | TM2 | 1 | 0.0087% |
| p.V95I | Chr18:60372067 | Gtt/Att | rs13447328 | TM2 | 1 | 0.0087% |
| p.V103I<sup>3</sup> | Chr18:60372043 | Gtc/Atc | rs2229616 | TM2 | - | - |
| p.T112M | Chr18:60372015 | aCg/aTg | rs13447329 | ECL1 | 4 | 0.0349% |
| p.N123S<sup>2</sup> | Chr18:60371982 | aAt/aGt | rs761982475 | TM3 | 1 | 0.0087% |
| p.S127L | Chr18:60371970 | tCg/tTg | rs13447331 | TM3 | 1 | 0.0087% |
| p.I137T | Chr18:60371940 | aTt/aCt | rs151102515 | TM3 | 2 | 0.0173% |
| p.H158R | Chr18:60371877 | cAt/cGt | rs202081467 | ICL2 | 2 | 0.0173% |
| p.S180P | Chr18:60371812 | Tca/Cca | rs193922685 | TM4 | 1 | 0.0087% |
| p.F184L<sup>2</sup> | Chr18:60371800 | Ttc/Ctc | - | TM4 | 1 | 0.0087% |
| p.F202L | Chr18:60371744 | ttC/ttA | rs138281308 | TM5 | 2 | 0.0175% |
| p.M215I | Chr18:60371705 | atG/atA | rs768687497 | TM5 | 1 | 0.0087% |
| p.A227T<sup>2</sup> | Chr18:60371671 | Gct/Act | rs201736647 | ICL3 | 1 | 0.0087% |
| p.R236C | Chr18:60371644 | Cgc/Tgc | rs758426526 | ICL3 | 1 | 0.0087% |
| p.G238VfsX4<sup>2</sup> | Chr18:60371636<br>-60371637 | gGt/gt | - | TM6 | 1 | 0.0087% |
| p.N240S | Chr18:60371631 | aAt/aGt | rs202228712 | TM6 | 2 | 0.0175% |
| p.I251WfsX34 | Chr18:60371598<br>-60371600 | ctGAtt/cttt | rs13447339 | TM6 | 1 | 0.0087% |
| p.I251L<sup>3</sup> | Chr18:60371599 | Att/Ctt | rs52820871 | TM6 | - | - |
| p.G252S | Chr18:60371596 | Ggc/Agc | rs13447336 | TM6 | 1 | 0.0087% |
| p.V253I | Chr18:60371593 | Gtc/Atc | rs187152753 | TM6 | 2 | 0.0175% |
| p.C271Y | Chr18:60371538 | tGt/tAt | rs121913562 | ECL3 | 1 | 0.0087% |
| p.S295P | Chr18:60371467 | Tca/Cca | rs368264587 | TM7 | 1 | 0.0087% |
| p.G323V<sup>2</sup> | Chr18:60371382 | gGa/gTa | rs926626133 | C-term | 1 | 0.0087% |

ALSPAC = Avon Longitudinal Study of Parents and Children; BP = base-pair position; C-term = C-terminus; ECL = extracellular loop; MAF = minor allele frequency; N-term = N-terminus; TM = transmembrane.
<sup>1</sup>Some mutations listed were carried by multiple individuals.
<sup>2</sup>Previously uncharacterised functionally.
<sup>3</sup>The common variants, p.V103I and p.I251L, were excluded from the main analyses.

The single exon gene, *MC4R*, was sequenced using a novel, cost-effective high-throughput approach, which involved pooling DNA from 5993 unrelated participants of ALSPAC into 120 pools (see **Methods**). We established that this approach had a sensitivity equivalent to whole exome sequencing (WES) of each individual DNA sample and ∼90% specificity in detecting single heterozygous *MC4R* mutations in pools of up to 50 individual DNA samples (see **Methods**). In total, 29 different non-synonymous mutations in *MC4R* were identified during sequencing of the cohort, including two frameshift/premature stop mutations and 27 missense mutations (**Table 1**). Two of the missense mutations were the commonly occurring p.V103I and p.I251L variants. Sanger sequencing confirmed the presence of all rare *MC4R* mutations and that carriers were heterozygous.

### The impact of MC4R mutations on canonical cAMP signalling

*MC4R* transduces external stimuli through Gα_s_-mediated activation of adenylyl cyclase, resulting in the increase of cytoplasmic levels of cyclic adenosine monophosphate (cAMP). Of the 29 non-synonymous mutations that were detected, 22 had previously been reported in terms of their ability to generate a cAMP response to melanocortin ligands and their association with human obesity (**Supplementary Table 2**). Of the 22 historically studied variants, two were reported to show complete loss of function (cLoF), nine to have a partial loss of function (pLoF), two to show gain of function (GoF) and nine to show wild-type (WT) like activity (see **Methods** and **Supplementary Table 2** for classification criteria and references).

**Table 2.** Prevalence estimates of *MC4R* LoF of cAMP accumulation identified in the ALSPAC sample used for analyses

| **Mutational characterisation** | **Frequency** | **Prevalence estimate**<br>**(% of total)** |
| --- | --- | --- |
| Synonymous, common variations or no LoF mutation | 5684 | 99.30 |
| GoF | 2 | 0.03 |
| WT-like | 21 | 0.37 |
| pLoF | 13 | 0.23 |
| cLoF | 4 | 0.07 |
| **Total** | **5724** | **100** |

*ALSPAC = Avon Longitudinal Study of Parents and Children; cLoF = complete loss of function; GoF = gain of function; LoF = loss of function; pLoF = partial loss of function; SD = standard deviation; WT = wild-type.*

We generated the seven previously uncharacterised mutants by site-directed mutagenesis and, in transiently transfected COS-7 cells, measured cAMP accumulation in response to escalating doses of [Nle^4^,D-Phe^7^]-α-melanocyte-stimulating hormone (NDP-αMSH) (**Figure 1**). Of the seven variants characterised, two were cLoF mutations (p.S85I and p.G238VfsX4) and one was a pLoF mutation (p.F184L). The four remaining variants (i.e., p.T5N, p.N123S, p.A227T and p.G323V) all displayed ‘WT-like’ activity (**Figure 1**). In total, there were 14 rare *MC4R* LoF mutations (four cLoF and 10 pLoF) identified in the study cohort (**Supplementary Table 2, Supplementary Table 3**).

**Table 3.**
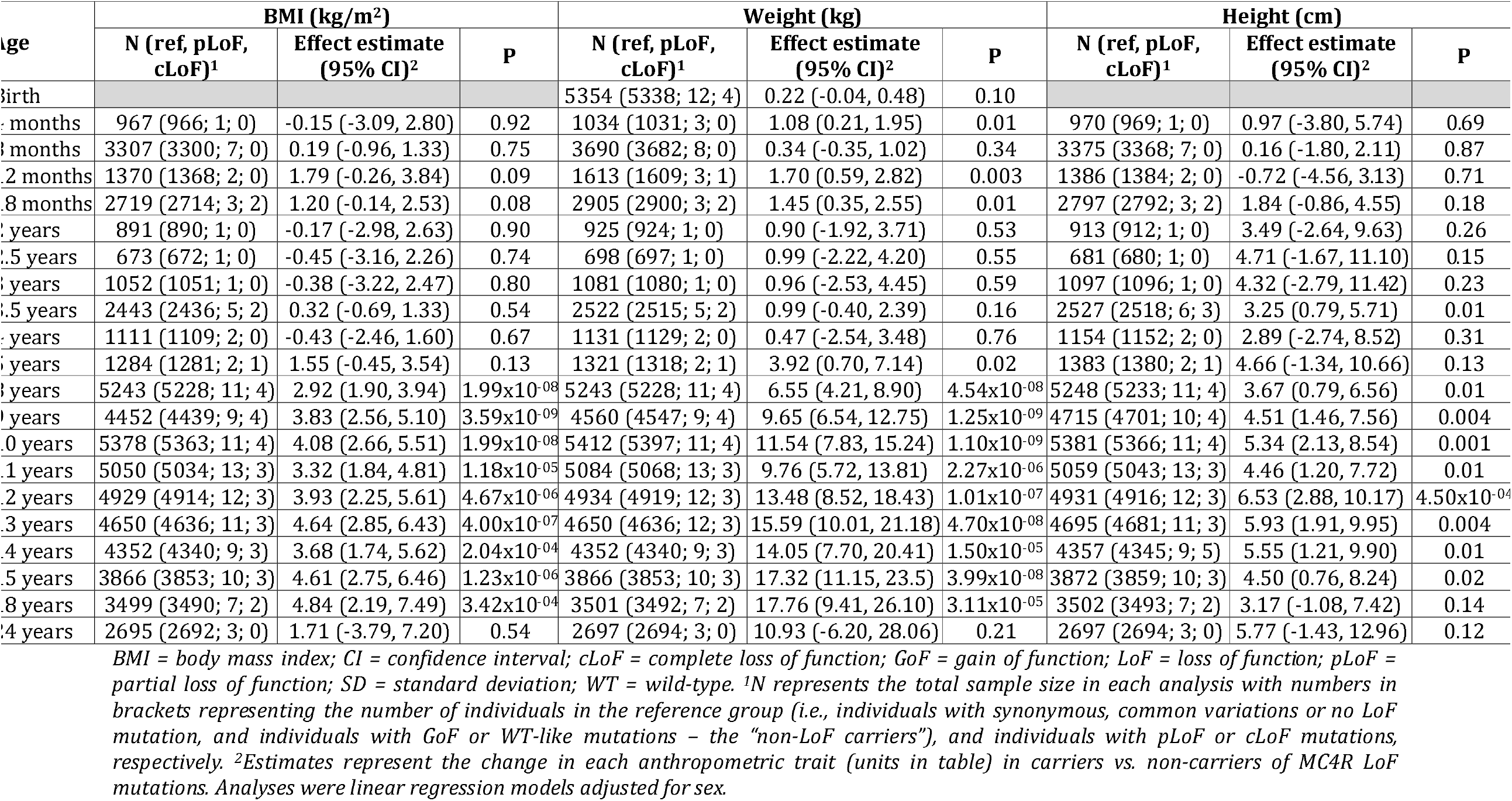
Age-specific associations between *MC4R* LoF of cAMP accumulation and BMI, weight and height

**Figure 1.**
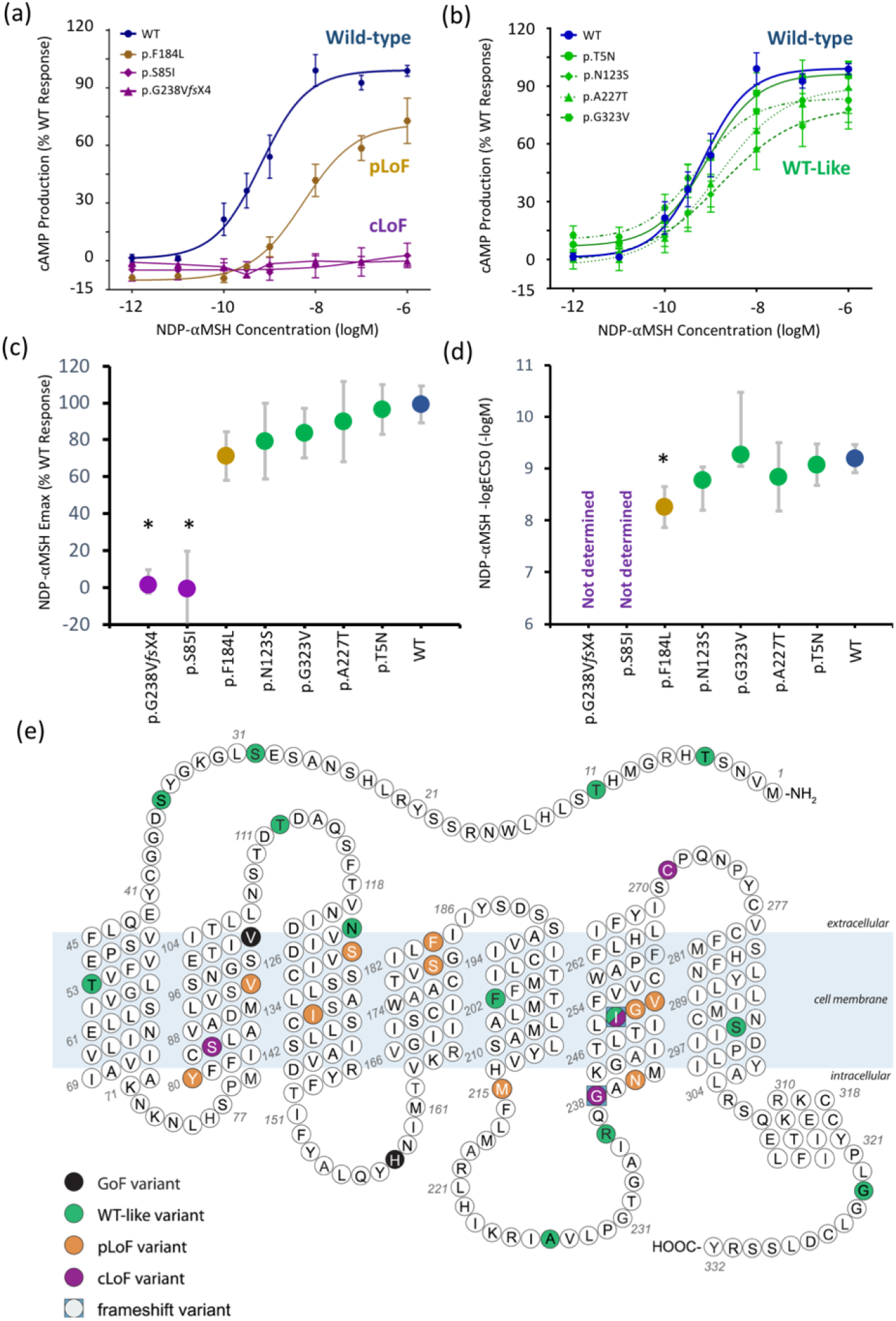
Ligand-activated cAMP accumulation of previously uncharacterized *MC4R* mutations. cAMP = cyclic adenosine monophosphate; cLoF = complete loss of function; GoF = gain of function; NDP-αMSH = [Nle4,D-Phe7]-α-melanocyte-stimulating hormone; pLoF = partial loss of function; WT-like = wild-type like; E_max_ = Relative maximal efficacy; EC50 = Half maximal effective concentration (a) Dose-response characteristics of NDP-αMSH-mediated cAMP production for the previously uncharacterized loss of function mutations found in ALSPAC; (b) dose-response characteristics of the previously uncharacterised ‘WT-like’ variants; (c) Relative maximal efficacy(E_max_) of NDP-MSH on MC4R mutants compared to WT, represented as estimated % WT response +/-95% CI; (d) The potency of NDP-MSH (– logEC50) on MC4R mutants, represented as estimated –logEC50 +/-95% CI; (e) schematic representation of MC4R showing all 29 mutations identified in the study cohort and their functional classification. There are 2 mutations at the p.I251 residue, p.I251L (WT-like) and p.I251WfsX34 (cLoF). *p<0.05

More recently, β-arrestin-2 coupling has been postulated to provide an important alternative post-receptor signal relevant to the control of body weight^25^. We examined NDP-αMSH-induced β-arrestin-2 coupling for all 27 rare variants (the common variants p.V103I and p.I251L were excluded) in transiently transfected HEK-293 cells. Using the same efficacy (E_max_) and potency (EC_50_) based criteria, we found that 10 of the 14 variants that were annotated as LoF for cAMP accumulation also showed impaired β-arrestin-2 coupling (**Supplementary Figure 1, Supplementary Table 3**). As cAMP is still considered to be the canonical signalling pathway for *MC4R*, our primary analyses of the association between *in vitro* function and clinical phenotype were undertaken using the cAMP-based functional classification with β-arrestin-2-based functional classification as a sensitivity analysis.

### Identification of rare variant carriers

Once we completed the functional characterisation of *MC4R* mutations, we unencrypted the sequenced pools to identify specific individuals carrying these mutations in ALSPAC. Of the 5993 individuals sequenced, a total of 5724 were used in the following analyses characterising the prevalence and downstream effects of *MC4R* LoF mutations on anthropometric traits due to exclusions of duplications and related individuals (see **Methods** for QC process). Of these 5724 participants, 40 individuals carrying 27 unique variants were confirmed as true positives (see **Methods**). Of these, 17 individuals carried a heterozygous copy of one of the 14 LoF mutations (**Table 2**), giving a prevalence of 0.30% of LoF mutations (i.e., approximately 1 in 337). Four participants (0.07%) carried a cLoF mutation and 13 (0.23%) carried a pLoF mutation. Twenty-one (0.37%) individuals carried WT-like mutations and two (0.03%) individuals had GoF mutations.

### Age-specific associations between MC4R mutations and anthropometric traits

Age-specific analyses were conducted using linear regression across all measures of selected anthropometric traits between birth and 24 years. There was a positive association between carriage of *MC4R* LoF mutations and BMI in childhood, adolescence and adulthood, with the mean difference increasing over time from as early as 5 years. This effect was greatest at age 18 years (**Table 3, Figure 2**), where the mean difference in BMI between carriers and non-LoF carriers of *MC4R* mutations was 4.84kg/m^2^ (95% CI: 2.19, 7.49; P=3.42×10^−04^). Similarly, there was a positive association between carriage of *MC4R* LoF mutations and weight (**Table 3, Supplementary Figure 2**), with the greatest difference between carriers and non-LoF carriers seen at 18 years (mean difference: 17.76kg; 95% CI: 9.41, 26.10; P=3.11×10^−05^). There was a smaller overall effect of *MC4R* LoF carriage on height over time (**Table 3, Supplementary Figure 3**), with the greatest difference at 12 years (mean difference: 6.53cm; 95% CI: 2.88, 8.54; P=4.50×10^−04^).

**Figure 2.**
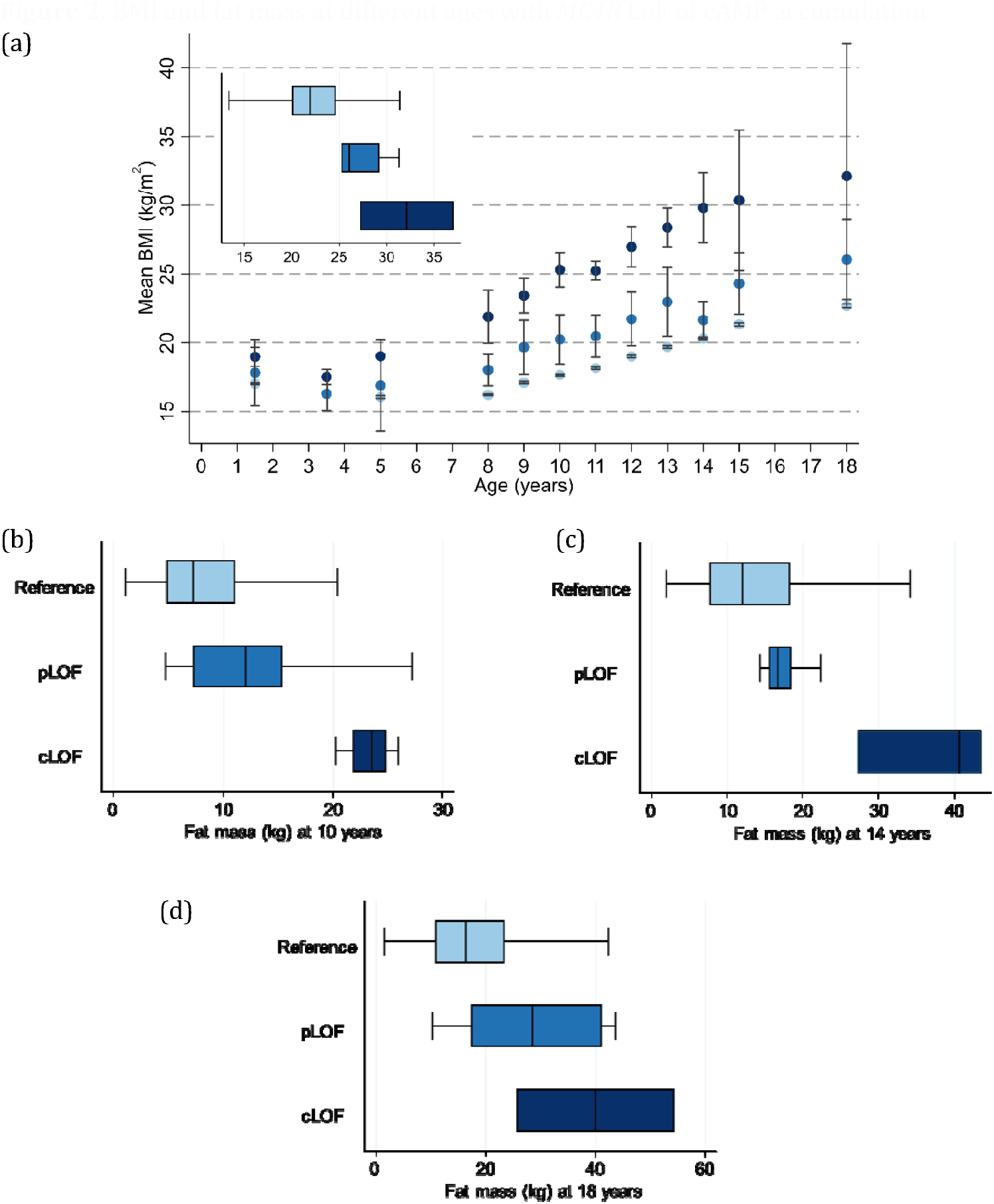
BMI at different ages with *MC4R* LoF of cAMP accumulation. BMI = body mass index; cLoF = complete loss of function; GoF = gain of function; LoF = loss of function; pLoF = partial loss of function; WT-like = wild-type like. (a) Mean BMI at different ages and (b) box plot showing distribution of BMI at age 18 years (exemplar) with MC4R LoF of cAMP (carriers of pLoF and cLoF) and the reference group (i.e., non-LoF carriers – combining individuals with synonymous, common variations or no LoF mutation and those with WT-like and GoF mutations). Figures only show results where all LoF mutations (i.e., pLoF and cLoF mutations) were represented by at least one individual at all time points between birth and 24 years.

**Figure 3.**
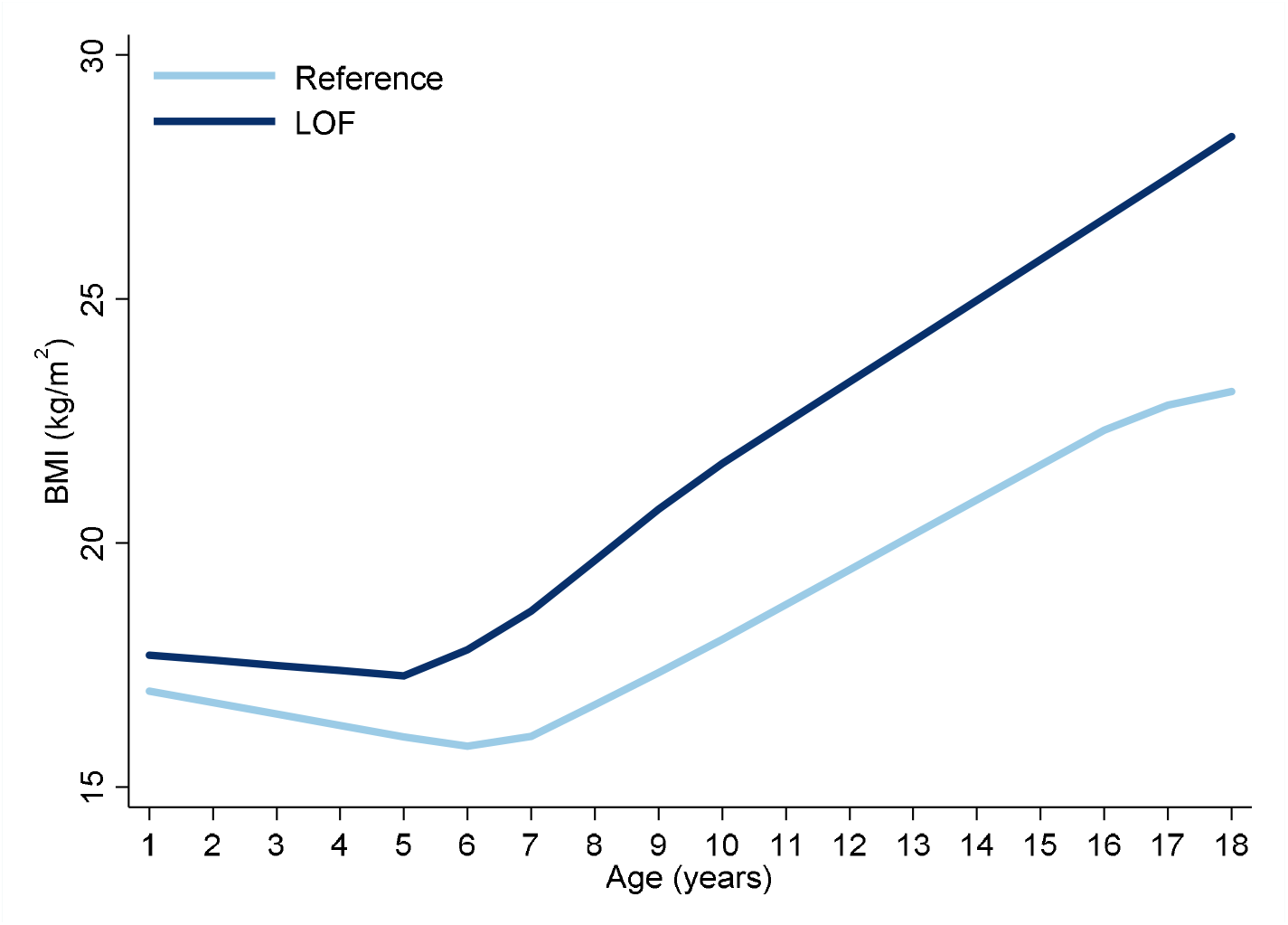
Association between *MC4R* LoF of cAMP accumulation with BMI trajectory between the ages of 18 months and 18 years using linear spline multi-level models. cLoF = complete loss of function; GoF = gain of function; LoF = loss of function; pLoF = partial loss of function; WT-like = wild-type like. Values for the reference group (i.e., all individuals with synonymous, common variations or no LoF mutation and those with WT-like and GoF mutations) and LoF mutations (i.e., combining pLoF and cLoF mutations) are depicted in light and dark blue, respectively. Estimates and confidence intervals of these associations can be seen in Supplementary Table 10.

*MC4R* LoF also showed a positive association with fat mass measured by dual energy x-ray absorptiometry (DXA) (**Supplementary Table 4**), with the greatest difference between carriers and non-LoF carriers at 18 years (mean difference: 14.78kg; 95% CI: 8.56, 20.99; P=3.27×10^−06^). The positive association between *MC4R* LoF and lean mass was also consistent over time (**Supplementary Table 4**), with the greatest difference between carriers and non-LoF carriers seen at 12 years (mean difference: 4.28kg; 95% CI: 2.08, 6.48; P=1.38×10^−04^).

Of the four waist-hip ratio (WHR) measures available in ALSPAC, the mean difference in WHR with *MC4R* LoF was the same at ages 10, 12 and 24 years, with carriers of *MC4R* LoF mutations increasing WHR by 0.04 (**Supplementary Table 4**) compared to non-LoF carriers. This difference was smaller at age 8 years (0.01; 95% CI: -0.01, 0.03; P=0.48) and increased to 0.04 at 24 years (95% CI: -0.03, 0.10; P=0.29); however, there were no individuals carrying a cLoF mutation and a measure of WHR at age 24 years.

In contrast to the associations seen between *MC4R* LoF carriage and anthropometric traits, there were no substantive differences in BMI, weight or height among individuals carrying “WT-like” receptors compared to non-LoF carriers not carrying WT-like mutations (**Supplementary Table 5**).

Previous studies have reported that carriers of *MC4R* LoF mutations have somewhat lower blood pressure (BP) than equally obese people who are WT at *MC4R*^26^. In this study, there was evidence that *MC4R* LoF mutation carriers had slightly higher systolic blood pressure (SBP) and left ventricular mass index (LVMI) but almost no difference in diastolic blood pressure (DBP) and central BP compared to non-LoF carriers between 8 and 18 years (**Supplementary Figure 4, Supplementary Figure 5**). These differences largely attenuated (or, indeed reversed) when adjusting for BMI at the same age. Of note, however, between ages 10 and 12 years, carriers of the *MC4R* LoF mutations had a DBP that was ∼3-5mmHg lower after correction for BMI and sex.

**Figure 4.**
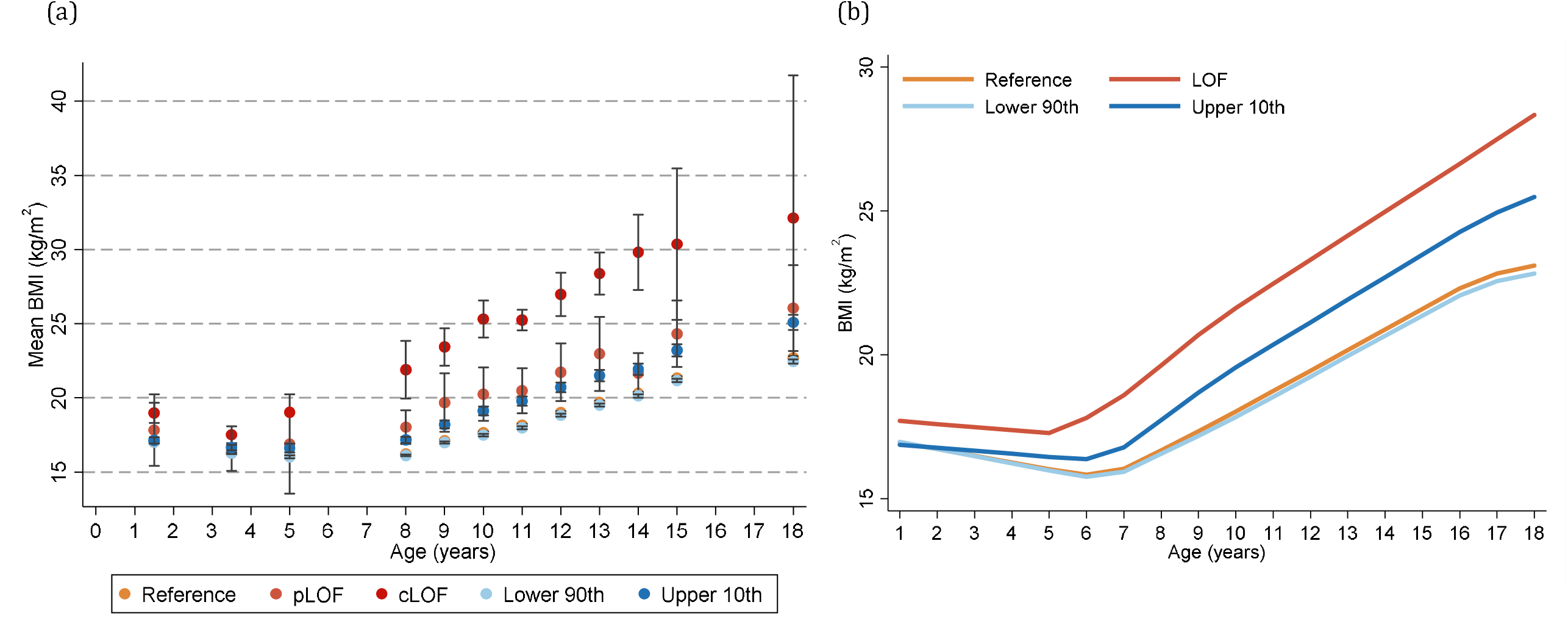
Comparison of the age-specific and longitudinal associations of *MC4R* LoF of cAMP accumulation and a weighted genome-wide polygenic risk score with BMI. BMI = body mass index; cLoF = complete loss of function; GoF = gain of function; LoF = loss of function; pLoF = partial loss of function; WT-like = wild-type-like. (a) Mean BMI across ages and (b) BMI trajectory between the ages of 18 months and 18 years using linear spline multi-level models, with estimates and confidence intervals of these associations available in Supplementary Table 10 and Supplementary Table 16, comparing MC4R LoF mutations and weighted genome-wide polygenic risk score. Figures only show results where all LoF mutations for MC4R (i.e., pLoF and cLoF mutations) were represented by at least one individual at all time points between birth and 24 years for comparison with the weighted genome-wide polygenic risk score. The reference group included individuals with synonymous, common variations or no LoF mutation and those with WT-like and GoF mutations.

### Associations between MC4R mutations and anthropometric traits assessed longitudinally

Longitudinal analyses were conducted to examine the association between the *MC4R* LoF mutations and the trajectory of BMI, weight and height. Multi-level linear-spline models used to examine longitudinal associations between *MC4R* LoF and anthropometric traits performed well when predicting each trait (**Supplementary Tables 6**-**8**).

There was little evidence to suggest that birth weight (mean: 3.44kg) was related to *MC4R* LoF (**Supplementary Table 9**). The first measure (i.e., intercept of the linear-spline multi-level models) of BMI and height was at 18 months (mean: 16.84kg/m^2^ and 81.90cm, respectively) and there was little evidence that *MC4R* LoF was associated with a difference in either BMI or height at this age in ALSPAC (**Supplementary Table 10, Supplementary Table 11**).

The effect estimates for the mean difference in BMI change (kg/m^2^ per year in carriers vs. non-carriers of *MC4R* LoF mutations) between 18 months and 18 years were non-zero and consistently positive between the 18 months and 15 years, consistent with the age-specific analyses (**Figure 3, Supplementary Table 10**). Similarly, the effect estimates for the mean differences in weight change (kg per year) between birth and 18 years were consistently positive, consistent with the age-specific analyses (**Supplementary Figure 6, Supplementary Table 9**). There was comparatively stronger evidence for a consistently positive effect of *MC4R* LoF on weight change between the ages of 12 months to 8 years (0.84kg/year; 95% CI: 0.40, 1.28; P=1.63×10^−04^) and 8 and 15 years (1.33kg/year; 95% CI: 0.66, 1.99; P=9.62×10^−05^).

There was a modest increase in height with the *MC4R* mutation across childhood and adolescence, with an inverse association in adulthood; however, most confidence intervals for these differences overlapped the null (**Supplementary Figure 7, Supplementary Table 11**).

### Sensitivity analyses

#### Comparison with β-arrestin-2 coupling

The phenotypic associations of *MC4R* LoF status were largely similar independent of LoF status being defined by β-arrestin-2 coupling or cAMP accumulation assay. There was a consistently positive trend between *MC4R* LoF of β-arrestin-2 coupling BMI from age 3.5 years, and weight and height across the lifecourse (**Supplementary Table 12, Supplementary Figures 8, 9** and **10**, respectively), where effect estimates were consistently larger with impairment of β-arrestin-2 coupling from approximately 8 years than cAMP accumulation. Associations between β-arrestin-2 coupling-based classification and fat mass, lean mass and WHR were consistently positive across all time points and had larger (or, with WHR, comparable) effects than those derived with cAMP signalling data (**Supplementary Table 13**). However, it is worth noting that all confidence intervals of associations of impairment in cAMP accumulation and β-arrestin-2 coupling assays with anthropometric traits overlapped.

#### Comparison between rare and common variation

Given the considerable recent interest in the use of genome-wide polygenic risk score (PRS) to predict the development of obesity^27^, we compared the impact of carriage of LoF mutations in *MC4R* with the PRS developed by Khera et al.^27^ (comparing upper 10^th^ to lower 90^th^ percentile of the PRS distribution). The magnitude of the effect estimates of *MC4R* LoF on BMI between the ages of 3 and 18 years was approximately double that of obtained by the PRS (**Figure 4a, Supplementary Table 14**). Similarly, using multi-level linear-spline models, the effect sizes of the change in BMI at 18 months and between 18 months and 18 years were almost always larger with *MC4R* LoF mutation compared to the PRS (**Figure 4b, Supplementary Table 15, Supplementary Table 16**). Findings from the main analyses of impact of the *MC4R* LoF mutations on BMI were also persistent, albeit slightly attenuated, even when adjusting for the genome-wide PRS (**Supplementary Table 17**). Unsurprisingly, given the relative rarity of *MC4R* LoF compared to the common SNPs comprising the PRS, the latter explained more of the population variance in BMI than the former (e.g., 0.40% and 10% explained by *MC4R* LoF mutations and the PRS, respectively, in BMI at age 18 years).

## DISCUSSION

By studying a large, representative birth cohort in which anthropometric measures are available throughout childhood, adolescence and early adulthood, we have been able to provide estimates of the prevalence of functionally impaired mutations in the *MC4R* gene. In addition to this, we have provided estimates of the phenotypic impact of these mutations during growth and development. We find that mutations at *MC4R* are more frequent and have a consistent and sizeable association with adiposity compared to what has been suggested of previously^15^.

To establish the prevalence of *MC4R* non-synonymous mutations in a specific population-based study, we developed an approach based on initial pooled high-throughput sequencing. We validated this methodology against WES of individual DNA samples and showed it to have very high sensitivity and specificity. This approach, which has considerable cost advantages, is applicable to the detection of rare, including private, mutations in any gene of interest in large populations.

We estimated a prevalence of heterozygous *MC4R* LoF mutations in ALSPAC birth cohort to be 0.30% (with 0.23% carrying pLoF and 0.07% carrying cLoF mutations). Given the well understood demographic characteristics of ALSPAC^22,23^ and notwithstanding ancestry-specific deviations in frequency, it is reasonable to suggest that as many as 1 in every 337 people in the UK may carry a heterozygous LoF mutation in the *MC4R* gene. These estimates are approximately double the previous report in an adult population-based cohort of European ancestry^15^ and currently approximates to 200,000 UK residents. In a more speculative approach, if this figure were to be true for populations worldwide (which is not inconsistent with published data^28-30^), up to 22 million people could carry a heterozygous LoF mutation in *MC4R*.

*MC4R* deficiency in mice results in an increase in both fat and lean mass^2^, with early reports suggesting that the same is true in humans^13^. Our results are consistent with these observations, with evidence to suggest a substantial impact of *MC4R* LoF carrier status on BMI, weight, fat mass and lean mass, which was detectable from as early as 5 years. For example, at age 18 years, carriage of an *MC4R* LoF mutation was associated with a 17.76kg greater body weight, a 4.84kg/m^2^ higher BMI and a 14.78kg greater fat mass, with 88.9% of carriers being overweight or obese at that age (compared to 20.5% of non-LoF carriers; data not shown). *MC4R* deficiency has also been reported to be associated with an increase in linear growth velocity attributable to hyperinsulinemia and the absence of the suppression of growth hormone levels that is usually seen in other forms of obesity^31^. Consistent with this, we observed a trend towards increased height with *MC4R* LoF during longitudinal follow-up.

In a recent study of UK Biobank participants, Turcot et al. reported a somewhat smaller impact of carriage of a heterozygous nonsense mutation in *MC4R* on body weight and the prevalence of obesity, where the majority of carriers were not obese^19^. Whilst we similarly found that not all carriers were, indeed, obese (in fact, the distributions of carriers and non-carriers of *MC4R* LoF mutations overlapped in most cases), the impact of carriage of such LoF mutations was, as described, more substantial. However, these previous findings by Turcot et al. should be viewed in the light of the known selection bias in UK Biobank^32^, whose participants are on average lighter and healthier than unselected members of the UK population and the fact that the particular subset of UK Biobank participants analysed in this study contained a sub-population that disproportionately represented heavy smokers.

In the current analyses, carriers of mutations that were functionally WT-like were practically indistinguishable from other non-LoF carriers or carriers of GoF mutations in their anthropometric characteristics. This emphasises the importance of knowing the functional impact of any non-synonymous mutation found during diagnostic testing in obesity. Indeed, databases collating information on the likely pathogenicity of all known mutations (https://www.mc4r.org.uk/) are very helpful to clinicians in this regard but, until every possible mutation has been systematically generated and characterised as has been undertaken with PPARG^33^, for example, such databases will remain incomplete. The mutations found in the present study had a largely similar impact on β-arrestin-2 coupling and cAMP accumulation (and, thus, had a largely similar impact on anthropometric traits) and do not, therefore, contribute to addressing questions around the relevance of biased signalling.

We previously reported that obese patients with *MC4R* LoF mutations have lower BP compared to similarly obese non-LoF carriers^26^. In this study, while there was a trend for an inverse association between *MC4R* LoF and BMI-adjusted measurements of both arterial and central cardiovascular health, there was no clear indication of lower BP in the *MC4R* carriers across the lifecourse. This finding, albeit restricted to a limited age range and sample size, is consistent with previous observations regarding the cardiovascular effects of *MC4R* functional impairment^26^.

Genome-wide PRSs associated with BMI have recently received considerable attention as possible predictors of phenotypes such as obesity^27^. In that context, it is notable that the impact of carrying a functionally impaired *MC4R* locus on BMI was approximately double that of the common PRS used previously (comparing the lower 90^th^ and upper 10^th^ percentiles of the continuous PRS distribution). Indeed, this was seen to be an effect which was persistent after having adjusted for PRS. This observation is not incompatible with the possibility of a buffer or enhancer-effect being present as a result of the individual-level combination of rare genetic changes and PRS value^34^. However, results here do suggest that the rare changes at *MC4R* are likely to have a larger impact than more subtle and continuous on-average differences delivered by theoretically additive PRS contributions at an individual level - a contrast to the nature of effect when considering total population variance explained.

A particular advantage of ALSPAC is the availability of robust longitudinal phenotyping data throughout childhood, adolescence, and adulthood. Childhood obesity is strongly associated with adverse cardiometabolic outcomes in later life^35^. However, it appears that the long-term adverse health consequences of childhood obesity are driven by the tendency of the obese phenotype to persist into adult life^36,37^. While it is conceivable that rescue of the phenotype at this stage of development would reduce cardio-metabolic risk (this also suggested in our analyses adjusting for BMI), we know from a longitudinal assessment of adult *MC4R* mutation carriers, that penetrance of the phenotype increases with age^18^. It therefore seems likely that the obese phenotype of *MC4R* LoF mutation carriers in ALSPAC cohort will persist or may even worsen with age.

Limitations of our study include the relatively small number of individuals carrying *MC4R* LoF mutations found in the ∼6000 ALSPAC participants sequenced. This led to wide confidence intervals for the analyses of the association with anthropometric traits. With this, and of importance to the external validity of our work, it is important to acknowledge the limited representation that ALSPAC offers to all other populations, especially those that are predominantly of European descent. Indeed, no single birth cohort, no matter how comprehensively collected, can be assumed to be representative of the nation or, indeed, the world.

That said, our study suggests that *MC4R* LoF mutations contribute substantially to adiposity traits, with effects starting in early childhood and persisting into adult life. In fact, heterozygous mutations that substantially impair the function of the *MC4R* gene may very well be found in several millions of people worldwide and will tend to increase the body weight and adiposity from an early age and persist across the lifecourse. Given the established association between *MC4R* LoF mutations and the complications of obesity such as type 2 diabetes and coronary artery disease^25^, this is of substantial clinical importance to the long-term health of individual carriers who will, on average, likely enter adult life carrying ∼15kg of extra fat mass. With a prevalence of ∼1/340, Melanocortin 4 receptor deficiency can no longer be considered a “rare disease”, the definition of which, in the UK, is a prevalence of <1/2000 and, in the US, is <200,000 affected patients nationally. If the prevalence in the USA reflects that found in the UK, we would predict that there are around a million Americans whose weight is substantially increased by the carriage of an *MC4R* mutation. Efforts to reduce obesity and maintain a healthy weight in carriers of *MC4R* LoF mutations, through diet and physical activity will likely need to begin early in life and be targeted in nature, to have an optimal chance of reducing the risks of developing obesity later in life. Pharmacological enhancement of residual intact melanocortin signalling could provide a clinically useful complement to such measures in these patients. The likely size of the population affected should help to stimulate investment in such therapeutic approaches.

## Data Availability

Full details of the cohort and study design have been described previously and are available at http://www.alspac.bris.ac.uk. Please note that the study website contains details of all the data that is available through a fully searchable data dictionary and variable search tool (http://www.bristol.ac.uk/alspac/researchers/our-data/). ALSPAC data are available through a system of managed open access. Data for this project was accessed under the project number B2891. The application steps for ALSPAC data access are highlighted below.
1. Please read the ALSPAC access policy which describes the process of accessing the data in detail, and outlines the costs associated with doing so.
2. You may also find it useful to browse the fully searchable research proposals database, which lists all research projects that have been approved since April 2011.
3. Please submit your research proposal for consideration by the ALSPAC Executive Committee.
You will receive a response within 10 working days to advise you whether your proposal has been approved. If you have any questions about accessing data, please.

## ACKNOWLEDGEMENTS

We are extremely grateful to all the families who took part in this study, the midwives for their help in recruiting them, and the whole ALSPAC team, which includes interviewers, computer and laboratory technicians, clerical workers, research scientists, volunteers, managers, receptionists and nurses. We acknowledge the technical support of Tolulope Osunnuyi and the NIHR BRC-MRC BioRepository at Cambridge Biomedical Research Centre. We also thank staff members of the Wellcome-Trust-MRC Institute of Metabolic Science (IMS) Genomics and Transcriptomic core and the Genomics Core at the CRUK Cambridge Institute for their experimental support for next-generation sequencing. Whole exome sequencing was obtained from ALSPAC (under the proposal numbered B2680) for comparison to the current study and we would like to thank Elise Robinson and Benjamin Neale from the BROAD institute for their contribution to the exome sequencing.

## FUNDING

The UK Medical Research Council and Wellcome (Grant ref: 102215/2/13/2) and the University of Bristol provide core support for ALSPAC. Genome-wide association data was generated by Sample Logistics and Genotyping Facilities at Wellcome Sanger Institute and LabCorp (Laboratory Corporation of America) using support from 23andMe. Mutational screening, sequencing and functional analyses were supported by MRC Metabolic Diseases Unit funding (MC_UU_00014/1). This publication is the work of all authors and KHW, NJT and SO serve as guarantors for the contents of this paper. KHW was supported by the Elizabeth Blackwell Institute for Health Research, University of Bristol and the Wellcome Trust Institutional Strategic Support Fund [204813/Z/16/Z]. NJT is a Wellcome Trust Investigator (202802/Z/16/Z), a work-package lead in the Integrative Cancer Epidemiology Programme (ICEP) that is supported by a Cancer Research UK programme grant (C18281/A19169) and works within the University of Bristol NIHR Biomedical Research Centre (BRC). DAH and LJC are supported by NJT’s Wellcome Trust Investigator grant (202802/Z/16/Z) and work within the Medical Research Council Integrative Epidemiology Unit [MC_UU_00011]. BYHL is supported by a BBSRC Project Grant (BB/S017593/1). AM holds a PhD studentship supported jointly by the University of Cambridge Experimental Medicine Training Initiative (EMI) programme in partnership with AstraZeneca (EMI-AZ). ISF was supported by the Wellcome Trust (098497/Z/12/Z), the NIHR Cambridge Biomedical Research Centre, the Botnar Fondation and the Bernard Wolfe Health Neuroscience Endowment and a Wellcome Developing Concept Fund award (with JM). SOR and GSHY is supported by the MRC Metabolic Disease Unit (MC_UU_00014/1) and SOR by a Wellcome Trust Investigator award (WT 095515/Z/11/Z) and NIHR Cambridge Biomedical Research Centre. The Wellcome-MRC Institute of Metabolic Science Genomics and transcriptomics core facility is supported by the Medical Research Council [MC_UU_00014/5] and the Wellcome Trust [208363/Z/17/Z].

## Authors contributions

KHW, BYHL, AM, GSHY, NJT and SOR designed the study. CZ, KR and KD conducted the genomic sequencing. JM and ISF contributed to the design of in vitro assays. WP, JHC, KL, KD, JM and AW planned and performed the in vitro studies. BYHL and AM conducted bioinformatic analysis and analysed the data from in vitro experiments. KHW conducted the analysis of phenotype data in the ALSPAC cohort. LJC critically reviewed the analysis in ALSPAC and the manuscript. DAH, KN and SN were involved in early conversations on the project and provided access to phenotypic, genotypic and exome data. KHW, BYHL, AM, GSHY, NJT and SOR wrote the manuscript and it was reviewed by all authors.

## Competing Interests statement

SOR has undertaken remunerated consultancy work for Pfizer, AstraZeneca, GSK, and ERX Pharmaceuticals.

## METHODS

### Study sample and measures

The Avon Longitudinal Study of Parents and Children (ALSPAC) is a large geographically-homogeneous prospective birth cohort from the southwest of England established to investigate environmental and genetic characteristics that influence health, development and growth of children and their parents^22,23,38^. Full details of the cohort and study design have been described previously and are available at http://www.alspac.bris.ac.uk. Please note that the study website contains details of all the data that is available through a fully searchable data dictionary and variable search tool (http://www.bristol.ac.uk/alspac/researchers/our-data/).

Briefly, 14541 pregnant women residing in the former county of Avon with an estimated delivery date of between the 1^st^ of April 1991 and the 31^st^ of December 1992 (inclusive) were enrolled to the study. Out of those initially enrolled, 13988 children who were alive at 1 year of age and have been followed up to date with measures obtained through regular questionnaires and clinical visits, providing information on a range of behavioural, lifestyle and biological data. When the oldest children were approximately 7 years of age, an attempt was made to bolster the initial sample with eligible cases who had failed to join the study originally. As a result, when considering variables collected from the age of seven onwards (and potentially abstracted from obstetric notes), there are data available for more than the 14541 pregnancies mentioned above. The number of new pregnancies not in the initial sample (known as Phase I enrolment) that are currently represented on the built files and reflecting enrolment status at the age of 24 is 913 (456, 262 and 195 recruited during Phases II, III and IV respectively), resulting in an additional 913 children being enrolled. The phases of enrolment are described in more detail in the cohort profile paper^22^.

The total sample size for analyses using any data collected after the age of seven is therefore 15454 pregnancies, resulting in 15589 foetuses. Of these 14901 were alive at 1 year of age. A 10% sample of the ALSPAC cohort, known as the Children in Focus (CiF) group, attended clinics at the University of Bristol at various time intervals between 4 to 61 months of age. The CiF group were chosen at random from the last 6 months of ALSPAC births (1432 families attended at least one clinic). Those excluded were mothers who had moved out of the area or were lost to follow-up, and those partaking in another study of infant development in Avon.

Ethical approval for the study was obtained from the ALSPAC Ethics and Law Committee and the Local Research Ethics Committees. Consent for biological samples has been collected in accordance with the Human Tissue Act (2004) and Informed consent for the use of data collected via questionnaires and clinics was obtained from participants following the recommendations of the ALSPAC Ethics and Law Committee at the time. Written informed consent was obtained from mothers at recruitment, from the main carers (usually the mothers) for assessments on the children from ages 7 to 16 years and, from age 16 years onwards, the children gave written informed consent at all assessments.

Academic attainment was derived by questionnaire asking whether the participant was still in full-time education (with possible answers of “yes” and “no”), when the participant was aged 18 years. Participant sex was measured from the birth notification as part of the cohort profile. Participant ethnicity was defined as either “White” or “Non-white” based on a questionnaire issued at approximately 32 weeks gestation completed by the participant’s mother. Household income was defined as the family income (in pounds) per week when the participant was 33 months old (defined as <£100, £100-199, £200-299, £300-399 or >£400). Age of mother at the birth of her first child was taken from a questionnaire administered during the 18-20 weeks gestational period of the ALSPAC participant. Maternal pre-pregnancy body mass index (BMI) was derived from weight (kg) and height (cm) measures obtained from a questionnaire administered during her pregnancy with the ALSPAC participant and calculated as weight divided by the square of height (kg/m^2^). Maternal weight gain was taken from obstetric records, calculated as the absolute weight gain from the last minus the first weight measurement (kg).

Highest household social class was a derived variable reflecting the highest social class based on occupation held by the participant’s mother or mother’s partner at 18 weeks gestation (with levels including “I – Professional”, “II – Managerial and technical”, “IIINM – Skilled non-manual”, “IIIM – Skilled manual”, “IV – Partly skilled” and “V – Unskilled”). Maternal and paternal education were derived from a questionnaire administered to the participant’s mother at 32 weeks gestation asking whether she and her partner had various qualifications, combined into a single variable reflecting her and her partner’s highest educational qualification (with levels including “CSE/none”, “Vocational”, “O-level”, “A-level” and “Degree”, where “CSE” is a Certificate of Secondary Education). Parity was defined as the number of previous pregnancies the participant’s mother had that resulted in either a live- or still-birth, obtained from a questionnaire administered at 18-20 weeks gestation.

Length and weight of each participant were measured at birth and at 4, 8, 12 and 18 months. Height (to the nearest millimetre) and weight (to the nearest 50g) were measured from 25 months to 24 years. For weight, the participant was encouraged to pass urine and undress to their underclothes. For height, children were positioned with their feet flat and heels together, standing straight so that their heels and shoulders came into contact with the vertical backboard. Equipment used (e.g., Harpenden Neonatometer or Stadiometer, Kiddimetre and Leicester measure for height and the Fereday 100kg combined scale, Soenhle scale, Seca scale and Tanita Body Fat Analyser for weight) for each measurement were comparable. In addition to the height and weight measures obtained at ALSPAC clinics, growth trajectories were carried out using linear spline multilevel modelling of height and weight from birth to when participants were 10 years. Therefore, any missing clinic values for height and weight were replaced with age-specific predicted values from growth trajectories^39^. BMI at all ages was calculated as weight or length (kg) divided by height (m) squared.

Both waist and hip circumferences were measured when the participants were a mean age of 8, 10, 12 and 24 years. Waist circumference was measured to the nearest millimetre at the minimum circumference of the abdomen between the iliac crests and the lowest ribs. Hip circumference was measured to the nearest millimetre at the point of maximum circumference around the participant’s hips. Waist-hip ratio (WHR) was calculated as the ratio of these two measurements.

Fat and lean masses (kg) were measured when participants were a mean age of 10, 12, 14, 15, 18 and 24 years using the Lunar prodigy narrow fan beam densitometer dual energy x-ray absorptiometry (DXA) scanner. The participant was asked to lie on the machine (in light clothing without any metal fastenings) and encouraged to keep as still as possible whilst the arm of the machine moved over, and two sources of X-ray scanned the participant.

Arterial blood pressure (BP) was measured when participants were a mean age of 3, 4, 5, 8, 10, 12, 13, 14, 15, 18 and 24 years old, with the appropriately sized cuff. Equipment used included Dinamap vital signs monitors (models 9300, 9301 and 8100) and Omron oscillometric devices (models MI-5, 705 IT, IntelliSense M6 and BP Cuff), which were comparable. Additionally, when participants were a mean age of 18 and 24 years, measures of cardiac structure and function were obtained. Of these measures, we used information about central BP and left ventricular mass index scaled by height to the power of 2.7 (LVMI, g/m^2.7^), as proxies for cardiovascular health^40^. Central BP was measured using radial artery tonometry with a SphygmoCor Px Pulse Wave Analysis System (Atcor Medical) at both age 18 and 24 years. Echocardiography was performed using a HDI 5000 ultrasound machine (Phillips) and P4-2 Phased Array ultrasound transducer (at age 18 years) and a Philips EPIQ 7G Ultrasound System (at age 24 years) using a standard examination protocol and left ventricular mass was estimated according to American Society of Echocardiography guidelines^41^.

Full details of all measures used in this study are available on the online dictionary: http://www.bristol.ac.uk/alspac/researchers/our-data/.

### Detection of *MC4R* mutations by pooled sequencing

#### Pooled high-throughput sequencing of MC4R

The pooled sequencing workflow is depicted in **Supplementary Figure 11a**. 20ng of 5993 DNA samples from ALSPAC were randomly combined into pools of 50 at the Medical Research Council Biorepository Unit. 10ng of pooled DNA was used for *MC4R* exon PCR with Q5 Hot Start High-Fidelity DNA Polymerase (NEB, Ipswitch MA, USA) and *MC4R* exon primers -27 bp upstream and +104 bp downstream of the protein coding region (**Supplementary Figure 11b, Supplementary Table 18**). The PCR product was purified using Agencourt Ampure XP beads (Beckman Coulter, Brea CA, USA), and quantified using the QuantiFlour dsDNA system (Promega, Wadison, WI, USA) and Tecan Infinite M1000 Pro plate reader (Männedorf, Switzerland). Sequencing libraries were constructed from 1ng of purified PCR product using the Nextera XT Library Preperation Kit with Nextera XT Index V2 barcodes (Illumina, San Diego, CA, USA) according to manufacturer’s instruction. The final libraries were purified using Agencourt Ampure Xp beads. Purified libraries were quantified by real-time quantitative PCR using the Kapa Library quantification kit (Roche, Basel, Switzerland) on a Quantstudio 7 Flex Real Time PCR instrument (ThermoFisher scientific, Waltham, MA, USA). Finally, the libraries were combined at 10nM for sequencing both ends for 150bp (PE150) on the Illumina HiSeq 4000 instrument at the CRUK Cambridge Institute Genomics Core. We achieved an even coverage throughout the protein coding region of *MC4R*, with a mean sequencing depth at 43,654 ± 356-fold per pool (**Supplementary Figure 11c**).

#### Sequencing bioinformatics

Sequence reads were mapped using BWA MEM algorithm (0.7.12) onto the Human GRCh38 (hg38) genome. PCR duplicates were removed using Picard 1.127 followed by indel realignment and base quality score recalibration using GATK 3.8 according to GATK Best Practices. The variant calls were generated by Varscan 2.4.2 mpileup2snp and mpileup2indel function with the following criteria: variant allele frequency (VAF) ≥0.05%, coverage ≥100, p-value <0.05 and strand filter set to ON. To maximise variant detection sensitivity, we started with an initial VAF cut-off at 0.5%, which was lower than the theoretically value of 1% in order to allow for technical errors and experimental bias, with an expectation of detecting false positives (FPs). The cut-off was readjusted using validation results from Sanger sequencing (see below).

#### Sanger sequencing for variant validation and rare variant carrier identification

Original DNA samples from all rare variant containing pools (except p.V103I and p.I251L) were retrieved for variant validation using traditional Sanger sequencing. The *MC4R* coding region was amplified, using GoTaq Green (Promega) Master Mix with 10ng DNA per 10µl PCR reaction and *MC4R* exon primers used in NGS (**Supplementary Table 18**). *MC4R* PCR cycling conditions were as follows: one cycle of Hot Start at 95^°^C for five minutes, then 35 cycles of the following: denaturation at 95^°^C for 30 seconds, annealing at 60^°^C for 30 seconds, extension at 72^°^C for 2 minutes. Then, one cycle of final extension at 72^°^C for five minutes.

Unincorporated primers and dNTPs were removed from the PCR reactions by digesting with Exonuclease I (Exo) (NEB) and Shrimp Alkaline Phosphatase (SAP) (NEB) as follows: 20 units of Exonuclease I and 1 unit of Shrimp Alkaline Phosphatase were added directly to the 10 µl PCR reaction; the EXO/SAP reaction was then incubated at 37^°^C for 20 minutes and then the enzymes were deactivated by incubating at 80^°^C for 15 minutes. This EXO/SAP PCR reaction was then used as the template for the Sanger Sequencing reaction.

Sanger sequencing reactions were set up using BigDye Terminator v3.1 Cycle Sequencing Kit (Thermal Fisher) in a 10µl reaction using 0.5µl of BigDye Terminator v3.1, 2µl 5x Sequencing buffer, 0.5µM sequencing primer and 1µl of the EXO/SAP PCR product which was made up to 10µl using Nuclease free water. The Sanger sequencing cycling conditions were as follows: denaturation at 95^°^C for 10 seconds, annealing at 50^°^C for five seconds, extension at 60^°^C for four minutes. This program was continued for 24 cycles in total.

Sanger sequencing reactions had unincorporated dye and primers and dNTPs removed using AxyPrep MAG PCR Clean-Up Kit (Axygen) according to manufacturer’s instructions. Purified sequencing products were resuspended in 30µl nuclease free water. Sanger sequencing reactions were analysed on a 3730 DNA Analyzer (Thermal Fisher). Sanger sequencing data files were analysed using Sequencher 4.8 Build 3767 (Gene Codes Corporation).

#### Specificity and sensitivity of pooled high-throughput sequencing

Excluding p.V103I and p.251L, the initial screen using a VAF 0.5% cut-off resulted in a total of 38 rare, non-synonymous *MC4R* variants with an estimated carriage of 57 individuals (data not shown). Of these, 40 individuals carrying 27 unique variants were confirmed as true positives (TP) by Sanger sequencing (**Supplementary Figure 11d**). The mean (± standard deviation, SD) VAF detected for TP was 1.18 (± 0.39%). We found a strong relationship between VAF and specificity, where all variants called at VAF<0.60% were FP (**Supplementary Figure 11d**), this indicates the likelihood of missing any true potential variants at VAF of <0.5% was extremely low. We also performed receiver operating curve (ROC) analysis and showed that VAF was a strong predictor for variant detection (AUC=0.976, **Supplementary Figure 11e**). Using findings from ROC, we adopted a final VAF cut-off at ≥0.60% and improved the specificity to 88.89%, whilst retaining all the 40 TP calls for downstream analysis.

To establish method sensitivity, we compared our *MC4R* variant call set with another ALSPAC whole-exome sequencing (WES) study of 2971 individuals. The *MC4R* locus in this study was sequenced at a depth of 28.15X. Within the overlap of 2451 (out of 5724) unique individuals sequenced in both studies (**Supplementary Figure 11f**), we found that the TP variant call sets were 100% concordant (**Supplementary Figure 11g**). This implies the sensitivity from our novel pooled sequencing method was on par with standard WES.

### Functional characterisation of *MC4R* mutations in vitro

#### cAMP accumulation assay

CV-1 in Origin with SV40 genes 7 (COS-7) cells were maintained in a growth medium containing low glucose Dulbecco’s modified eagle medium (Invitrogen, Carlsbad, CA, USA), 10% fetal bovine serum (Invitrogen), 1% Glutamax (Invitrogen), 100 U/ml penicillin and 100 mg/ml streptomycin (Sigma-Aldrich, IL, USA). COS-7 cells were kept at 37^°^C humidified air with 5% CO_2_.

Site-directed mutagenesis was performed on WT Human N-FLAG-*MC4R* PCDNA3.1(+) using Agilent QuikChange Lightning kit (Santa Clara, CA, USA) to generate all 7 previously uncharacterised *MC4R* variants for cAMP activity measurement. 30ng of plasmid carrying *MC4R* WT and variants were transfected into COS-7 cells using (Lipofectamine 2000, Invitrogen) for 24 hours. [Nle^4^,D-Phe^7^]-α-melanocyte-stimulating hormone (NDP-αMSH, Bachem, Bubendorf Switzerland), dissolved in 0.1% bovine serum albumin (BSA) and 1mM acetic acid at a stock concentration of 5mM, was added to cells at increasing final concentrations of 10^−12^ to 10^−6^M in the growth medium for 2 hours, before intracellular cAMP concentration measurement using a luminescence based HitHunter cAMP Assay for small molecules (Cat# DiscoverX 90-0075SM25 Eurofins DiscoverX, Fremont, CA, USA) and a Tecan Spark 10M microplate reader. The baseline and maximal luminescence signal was normalised to *MC4R* WT and a 4-point sigmoidal dose-response curve was fitted to normalised values from all replicates to determine the E_max_ and logEC_50_ using Graphpad Prism 7. Due to the lack of response, we did not perform a curve fit for complete LoF (cLoF) variants and only determined the relative E_max_ based on cAMP level measured at 10^−7^M NDP-αMSH.

#### *B*-arrestin-2 coupling assay

*To examine the* interactions between *MC4R* and β-arrestin-2, we used the NanoBiT protein/protein interaction assay (Promega). *MC4R* WT and variants were cloned into the pBiT1.1-C(TK/LgBiT) vector. 50ng of the *MC4R*-LgBiT and ARRB2-SmBiT were co-transfected into HEK293 cells as described by Lotta., et al. 2019^25^. Human embryonic kidney 293 (HEK-293) cells were maintained in high glucose Dulbecco’s modified eagle medium (Invitrogen), 10% fetal bovine serum (Invitrogen), 1% Glutamax (Invitrogen), 100 U/ml penicillin and 100 mg/ml streptomycin (Sigma-Aldrich, St Louis, MO, USA). HEK-293 cells were kept at 37^°^C humidified air with 5% CO_2_. 24 hours after transfection, culture medium was replaced with Opti-MEM I medium (Invitrogen) 30 minutes before luciferase activity was measured by the Tecan Spark 10M microplate reader set at 37^°^C and 5% CO_2_. After 2.5 minutes, 20µl of Nano-Glo Live Cell Assay System (Promega) was added and luciferase activity was measured for 10 minutes to generate the baseline signal. Cells were then stimulated with NDP-αMSH at 10^−12^ to 10^−6^M and luciferase activity was monitored for another 30 minutes. The area under the curve (AUC) above the baseline was then used to determine the coupling between *MC4R* and β-arrestin-2. For each individual experimental replicate, the AUC values were normalised to % maximum AUC of *MC4R* WT from the same experiment and a 3-point sigmoidal dose-response curve was fitted to determine E_max_ and logEC_50_. The average E_max_ and logEC_50_ values were used for LoF determination. The logEC_50_ was not used for cLoF mutants that exhibited no response. All calculations were performed with GraphPad Prism 6.

#### Identification of rare variant carriers

Once we completed the functional characterisation of *MC4R* mutations, we unencrypted sequenced pools to identify specific individuals carrying these mutations. This allowed the phenotypic characterisation of such functional impairment of *MC4R*. Of the 5993 individuals sequenced, five individuals had missing identifier information for linkage with the wider ALSPAC data set and 214 individuals were duplicated; therefore, these exclusions left 5774 participants in the sequenced set (note that none of the excluded individuals had an *MC4R* LoF mutation). After merging in all required clinic and questionnaire data from the ALSPAC cohort, there were related individuals (i.e., siblings) within the total sample. For appropriate comparisons between those included within and excluded from the sequenced set, all related individuals were excluded. Specifically, there were one set of quadruplets (none of which were in the sequenced set), four sets of triplets (one full set of which was in the sequenced set) and 255 sets of twins (48 complete sets of which were in the sequenced set). In addition to these 48 pairs of twins in the sequenced set, there were 35 sets of twins where one twin was in the sequenced set and the other twin was not in the sequenced set. To avoid removing as many individuals from the sequenced set as possible, the twin not in the sequenced set was removed in these instances (n=35).

A total of 216 individuals were excluded from the total sample to remove siblings (note that, at this point, none of these were from the sequenced set), which included 35 individuals (one of a pair of twins not in the sequenced set), three individuals from a quadruplet, two individuals each from three triplets (n=6), 172 individuals from twin sets. Then, siblings in the sequenced set were removed, which included the two individuals from the one triplet set and one individual from each of the 48 twins (n=50). After all exclusions, there were 5724 left in the sequenced set for all analyses.

To identify and estimate the prevalence of carriers of loss of function (LoF) mutations, tabulations were used, separated by gain of function (GoF), wild-type-like (WT-like), partial LoF (pLof) and cLoF mutations, in the sequenced set of ALSPAC participants.

### Statistical analyses

For this paper, we focused on *MC4R* LoF of cAMP production as our main analysis, with comparison to impairment of β-arrestin-2 coupling and to a genome-wide polygenic risk score (PRS) comprising over 2 million common genetic variants as sensitivity analyses. All analyses were conducted using Stata (version 15) and MLwiN version 3.04 called from Stata using the *runmlwin* command^42^.

#### Representation

To explore how representative the participants of the sequenced set were of the wider ALSPAC cohort, measures of education, socioeconomic status and parental factors were compared between individuals within the sequenced set and those not in the sequenced set. These variables were selected based on those used in previous papers comparing the ALSPAC cohort with statistics from a national sample of the United Kingdom^22,23^. These measures included academic attainment, sex, ethnicity, household income, maternal age at birth of first child, maternal pre-pregnancy BMI, maternal weight gain during pregnancy, highest household social class, parental education, and parity (see above for details on how these were measured).

Means and SDs of all continuous variables and percentages of binary or categorical variables were calculated and ttests were used to test whether summary statistics were different between participants included in the sequenced set and those in the wider ALSPAC cohort.

#### Age-specific associations between MC4R mutations and anthropometric traits

Age-specific analyses were conducted using linear regression across all available measures of BMI, height, weight, WHR, fat mass and lean mass between birth and 24 years. All individuals with data on the *MC4R* mutations, anthropometric trait and sex were included in each model (i.e., complete case analysis); therefore, sample sizes differ across ages.

*MC4R* LoF produced by the mutations was analysed by comparing carriers of LoF mutations (i.e., all individuals with pLoF and cLoF mutations) to “non-LoF carriers” (i.e., all individuals with synonymous or common variants and no LoF mutation and those carrying WT-like or GoF mutations) as the reference group. Effect estimates therefore represent the mean difference in each anthropometric trait in carriers vs. non-carriers of *MC4R* LoF mutations. Associations were adjusted only for participant sex. We also assessed the effect of carriage of WT-like mutations on BMI, weight, and height compared to those with no detected LoF variant.

As *MC4R* mutation carriers have previously been reported to have lower BP than equivalently obese WT individuals, we additionally examined the association between *MC4R* LoF (defined in the same way as above) on clinically relevant obesity-related traits. Age-specific analyses were therefore conducted with measures of arterial and central BP and LVMI across the lifecourse, as proxies for cardiovascular health, both in a sex-adjusted model and a model additionally adjusted for BMI measured at the same occasion.

We also plotted the mean anthropometric trait at each time point, separating out carriers to the component mutational parts (i.e., comparing the reference “non-LoF carrier” group to pLoF and cLoF separately). Not all individuals with *MC4R* mutations had anthropometric/cardiovascular measurements available at all time points between birth and 24 years and, on some occasions, all LoF groups were not represented (e.g., no individuals carrying a cLoF mutation had anthropometric data at age 24). In these instances, tables show results at all time points with all contributing individuals and figures only show results where all LoF mutational groups (pLoF and cLoF mutations) were represented by at least one individual.

Sensitivity analyses were also conducted to assess the effect of excluding individuals carrying GoF mutations in main analyses, leaving only individuals with no LoF mutations and those carrying WT-like mutations in the “non-LoF carriers” reference group, but this made very little difference to findings (results available on request).

#### Associations between MC4R mutations and anthropometric traits assessed longitudinally

Longitudinal analyses using linear-spline multi-level models were conducted to examine the association between the *MC4R* mutations and change in each anthropometric trait across the lifecourse. Given the limited number of WHR, fat mass and lean mass observations between birth and 24 years, longitudinal analyses focused on characterising the association between the *MC4R* mutations and BMI, weight, and height only. Additionally, given the lack of individuals carrying a cLoF mutation and anthropometric traits at age 24 years, longitudinal analyses were restricted to capture *MC4R*-driven anthropometric variation from the first instance that all LoF mutational groups were represented (i.e., 18 months for BMI and height, and birthweight) to 18 years of age. Multi-level models estimate the mean trajectories of each anthropometric trait, while accounting for non-independence of repeated measures within individuals, change in scale and variance of measures over time, and differences in the number and timing of measurements between individuals (using all available data from all eligible participants under a missing-at-random assumption). Linear splines allow knot points to be fitted at different ages to derive periods of change that are approximately linear. All participants with at least one measure of the anthropometric traits were included under a missing-at-random assumption to minimize selection bias in trajectories estimated using linear spline multi-level models (with two levels: measurement occasion and individual).

Knot points were placed as follows for each anthropometric trait based on the distribution and longitudinal pattern of measures between the earliest measure and 18 years: at ages 3.5, 5, 8 and 15 years for BMI; at ages 5 and 15 years for height; and at ages 1, 8 and 15 years for weight. Interaction terms between the variable indicating *MC4R* LoF (comprising the “non-LoF carrier” reference group (i.e., individuals with synonymous, common variations or no LoF mutation and individuals with GoF mutations) and carriers of LoF mutations) and each spline were included in the models to estimate the difference in the intercepts (earliest anthropometric trait measurement) and slopes (change in anthropometric trait from the earliest measure to 18 years across splines) between *MC4R* LoF. Additionally, interaction terms between sex and each spline were included to estimate the difference in intercepts and slopes between males and females; therefore, models were adjusted for sex.

### Sensitivity analyses

#### Comparison with β-arrestin-2 coupling

To understand the impact of β-arrestin-2-based *MC4R* functional classification on adiposity relative to those acting through cAMP signalling, we examined the age-specific associations between β-arrestin-2 LoF mutations with the same anthropometric traits as described above. Functional impairment of β-arrestin-2 coupling was coded in the same way as main analyses – comparing carriers (i.e., those carrying pLoF and cLoF mutations) to non-LoF carriers (i.e., individuals with synonymous, common variations or no LoF mutation and those with either WT-like or GoF mutations).

#### Comparison between rare and common variation

There has been growing interest in the use of the PRSs as predictors of phenotypes such as obesity and related metabolic fluctuations^27^. We assessed the comparative variation in anthropometric traits (namely, BMI) between carriage of *MC4R* LoF mutations with that of a genome-wide PRS comprising over 2 million common genetic variants weighted according to their effect sizes on BMI.

The weighted genome-wide PRS was generated in the same way as that used by Khera et al.^27^ and categorized as a binary variable reflecting individuals in the lower 90^th^ and upper 10^th^ percentiles of the PRS distribution. Age-specific analyses were conducted in the same way as above with all available measures of BMI and longitudinal models were performed on data capturing BMI from 18 months to 18 years of age, with groups of the PRS as the independent variable. For direct comparison with the effect sizes of the *MC4R* rare variation, these analyses were restricted to the individuals who were in the original sequenced set (n=5724).

To assess whether the genetic impact of the *MC4R* LoF mutations and the PRS were interconnected, we adjusted the main age-specific analyses of the association between *MC4R* LoF mutations and BMI for the genome-wide PRS at all available ages. We also calculated the variance in BMI explained by the PRS as compared to the *MC4R* LoF mutations using the measurement at age 18 years as an exemplar. We took the subset of participants who had both BMI measured at age 18 years and a derived PRS (N=3164, including 7 carriers) and ran a linear regression firstly with LoF mutation carrier status as the independent variable and secondly with the PRS as the independent variable (with BMI at age 18 years as the dependent variable in both cases). The R^2^ from these models was taken as an estimate of the variance explained in each case.

## Notes

### Competing Interest Statement

SOR has undertaken remunerated consultancy work for Pfizer, AstraZeneca, GSK, and ERX Pharmaceuticals. All other authors have declared no competing interests.

### Author Declarations

Ethical approval for the study was obtained from the ALSPAC Ethics and Law Committee and the Local Research Ethics Committees. Consent for biological samples has been collected in accordance with the Human Tissue Act (2004) and Informed consent for the use of data collected via questionnaires and clinics was obtained from participants following the recommendations of the ALSPAC Ethics and Law Committee at the time.

