## Supplementary Tables and Figures for "Prevalence and expressivity of loss of function mutations in the Melanocortin 4 Receptor (*MC4R*) in a UK birth cohort"

**Supplementary Table 1.** Differences in characteristics in those sequenced for *MC4R* LoF mutations and those not sequenced from the rest of ALSPAC

| **Trait** | **Included** | | **Excluded** | | **Difference** | |
| --- | --- | --- | --- | --- | --- | --- |
|  | **N** | **Mean (SD) or %** | **N** | **Mean (SD) or %** | **Mean (SD) or %** | **P-value^1^** |
| ***Parental characteristics*** | | | | | | |
| Maternal age at birth of first child | 5317 | 25.49 (4.84) | 7835 | 23.55 (4.95) | 1.94 (0.09) | 1.42x10^-109^ |
| Maternal pre-pregnancy BMI | 4893 | 22.91 (3.74) | 6621 | 22.95 (3.93) | -0.03 (0.07) | 0.63 |
| Maternal weight gain in pregnancy | 4919 | 12.63 (4.55) | 7282 | 12.51 (4.69) | 0.12 (0.09) | 0.16 |
| Parity | 5236 | 0.80 (0.92) | 7695 | 0.87 (1.06) | -0.07 (0.02) | 3.62x10^-05^ |
| Highest household social class | 5,137 |  | 7,124 |  |  | 2.08x10^-58^ |
| Professional (I) | 582 | 11.33 | 514 | 7.22 | 4.11 |  |
| Managerial and technical (II) | 2,161 | 42.07 | 2,398 | 33.66 | 8.41 |  |
| Skilled, non-manual (III) | 1,506 | 29.32 | 2,269 | 31.85 | -2.53 |  |
| Skilled, manual (III) | 556 | 10.82 | 1,119 | 15.71 | -4.89 |  |
| Partly skilled (IV) | 280 | 5.45 | 647 | 9.08 | -3.63 |  |
| Unskilled (V) | 52 | 1.01 | 177 | 2.48 | -1.47 |  |
| Family income (£/wk) | 4353 |  | 4365 |  |  | 1.58x10^-35^ |
| <100 | 249 | 5.72 | 516 | 11.82 | -6.10 |  |
| 100-199 | 657 | 15.09 | 882 | 20.21 | -5.12 |  |
| 200-299 | 1,256 | 28.85 | 1,217 | 27.88 | 0.97 |  |
| 300-399 | 1,023 | 23.50 | 827 | 18.95 | 4.55 |  |
| >400 | 1,168 | 26.83 | 923 | 21.15 | 5.68 |  |
| Maternal education at 33 months | 5,041 |  | 6,505 |  |  | 2.31x10^-79^ |
| CSE | 493 | 9.78 | 1,231 | 18.92 | -9.14 |  |
| Vocational | 426 | 8.45 | 787 | 12.10 | -3.65 |  |
| O-level | 1,836 | 36.42 | 2,427 | 37.31 | -0.89 |  |
| A-level | 1,401 | 27.79 | 1,359 | 20.89 | 6.90 |  |
| Degree | 885 | 17.56 | 701 | 10.78 | 6.78 |  |
| Paternal education at 33 months | 4,781 |  | 5,838 |  |  | 9.22x10^-27^ |
| CSE | 688 | 14.39 | 1,192 | 20.42 | -6.03 |  |
| Vocational | 407 | 8.51 | 594 | 10.17 | -1.66 |  |
| O-level | 1,123 | 23.49 | 1,390 | 23.81 | -0.32 |  |
| A-level | 1,426 | 29.83 | 1,651 | 28.28 | 1.55 |  |
| Degree | 1,137 | 23.78 | 1,011 | 17.32 | 6.46 |  |
| ***Participant characteristics*** | | | | | | |
| Continued education at 18 years (% yes) | 2902 | 88.15 | 1147 | 87.36 | 0.79 | 0.49 |
| Sex (% female) | 5717 | 48.52 | 13543 | 48.38 | 0.14 | 0.86 |
| Ethnicity (% white) | 5150 | 96.08 | 6820 | 94.11 | 1.97 | 1.04x10^-06^ |

*ALSPAC = Avon Longitudinal Study of Parents of Children; BMI = body mass index; CSE = Certificate of Secondary Education; SD = standard deviation.*

*^1^For continuous variables, P-values were derived from a ttest of the mean difference between those included vs. excluded from the group of sequenced individuals. For binary and ordered categorical variables, P-values were derived from a ttest of the mean difference in trend across all categories.*

**Supplementary Table 2.** Functional annotation of all *MC4R* mutations.

[please find in the accompanying Excel file]

**Supplementary Table 3.** List of all *MC4R* mutations grouped by their cAMP LoF classification and their corresponding β-arrestin-2 classification.

| **Mutation** | **cAMP classification** | **β-arrestin-2 classification** |
| --- | --- | --- |
| **p.S85I** | cLoF | cLoF |
| **p.G238V*fs*X4** | cLoF | cLoF |
| **p.I251W*fs*X40** | cLoF | cLoF |
| **p.C271Y** | cLoF | cLoF |
| **p.Y80C** | pLoF | cLoF |
| **p.V95I** | pLoF | pLoF |
| **p.S127L** | pLoF | pLoF |
| **p.I137T** | pLoF | pLoF |
| **p.S180P** | pLoF | pLoF |
| **p.V253I** | pLoF | pLoF |
| **p.F184L** | pLoF | WT-Like |
| **p.M215I** | pLoF | WT-Like |
| **p.N240S** | pLoF | GoF |
| **p.G252S** | pLoF | WT-Like |
| p.T5N | WT-like | WT-Like |
| p.S30F | WT-like | WT-Like |
| p.S36T | WT-like | WT-Like |
| p.T112M | WT-like | WT-Like |
| p.N123S | WT-like | WT-Like |
| p.F202L | WT-like | WT-Like |
| p.A227T | WT-like | WT-Like |
| p.R236C | WT-like | WT-Like |
| p.S295P | WT-like | pLoF |
| p.G323V | WT-like | WT-Like |
| p.T11A | WT-like | GoF |
| p.T53I | WT-like | GoF |
| p.H158R | GoF | GoF |

*cAMP = cyclic adenosine monophosphate; cLoF = complete loss of function; GoF = gain of function; LoF = loss of function; pLoF = partial loss of function; WT-like = wild-type like*. *Individuals with any of the 14 LoF mutations marked in bold were those that comprised the "carrier" group (i.e., combining both cLoF and pLoF mutations) in main analyses of the association between MC4R LoF and anthropometric traits. These were compared to the reference group (i.e., non-LoF carriers - comprising individuals with synonymous, common variations or no LoF mutation and individuals with any of the listed (non-bold) GoF or WT-like mutations).*

**Supplementary Table 4.** Age-specific associations between *MC4R* LoF of cAMP accumulation and WHR, fat mass and lean mass

| **Age** | **Fat mass (kg)** | | | **Lean mass(kg)** | | | **WHR** | | |
| --- | --- | --- | --- | --- | --- | --- | --- | --- | --- |
|  | **N (ref, pLoF, cLoF)^1^** | **Effect estimate (95% CI)^2^** | **P** | **N (ref, pLoF, cLoF)^1^** | **Effect estimate (95% CI)^2^** | **P** | **N (ref, pLoF, cLoF)^1^** | **Effect estimate (95% CI)^2^** | **P** |
| 8 years |  |  |  |  |  |  | 5078 (5063; 11; 4) | 0.01 (-0.01, 0.03) | 0.48 |
| 10 years | 5109 (5095; 10; 4) | 8.16 (5.59, 10.72) | 4.71x10^-10^ | 5109 (5095; 10; 4) | 3.24 (1.63, 4.85) | 8.06x10^-05^ | 5339 (5324; 11; 4) | 0.04 (0.02, 0.06) | 0.001 |
| 12 years | 4874 (4859; 12; 3) | 8.64 (5.33, 11.96) | 3.39x10^-07^ | 4874 (4859; 12; 3) | 4.28 (2.08, 6.48) | 1.38x10^-04^ | 4926 (4911; 12; 3) | 0.04 (0.01, 0.06) | 0.01 |
| 14 years | 4295 (4283; 9; 3) | 9.76 (5.49, 14.04) | 7.86x10^-06^ | 4295 (4283; 9; 3) | 3.81 (0.49, 7.14) | 0.02 |  |  |  |
| 15 years | 3750 (3737; 10; 3) | 12.34 (7.91, 16.78) | 5.27x10^-08^ | 3750 (3737; 10; 3) | 4.04 (1.04, 7.04) | 0.01 |  |  |  |
| 18 years | 3408 (3399; 7; 2) | 14.78 (8.56, 20.99) | 3.27x10^-06^ | 3408 (3399; 7; 2) | 2.00 (-1.48, 5.47) | 0.26 |  |  |  |
| 24 years | 2631 (2628; 3; 0) | 9.08 (-2.67, 20.84) | 0.13 | 2631 (2628; 3; 0) | 1.58 (-5.58, 8.74) | 0.67 | 2692 (2689; 3; 0) | 0.04 (-0.03, 0.10) | 0.29 |

*CI = confidence interval; cLoF = complete loss of function; GoF = gain of function; LoF = loss of function; pLoF = partial loss of function; SD = standard deviation; WHR = waist-hip ratio WT = wild-type.*

*^1^N represents the total sample size in each analysis with numbers in brackets representing the number of individuals in the reference group (i.e., individuals with synonymous, common variations or no LoF mutation and individuals with GoF or WT-like mutations – the “non-LoF carriers”), and individuals with pLoF or cLoF mutations, respectively. ^2^Estimates represent the change in each anthropometric trait (units in table) in carriers vs. non-carriers of MC4R LoF mutations. Analyses were linear regression models adjusted for sex.*

**Supplementary Table 5.** Age-specific associations showing mean differences in BMI, weight and height comparing individuals with WT-like mutations for cAMP accumulation and non-LoF carriers not carrying WT-like mutations.

| **Age** | **BMI (kg/m^2^)** | | | **Weight (kg)** | | | **Height (cm)** | | |
| --- | --- | --- | --- | --- | --- | --- | --- | --- | --- |
|  | **N (ref, WT-like)^1^** | **Effect estimate (95% CI)^2^** | **P** | **N (ref, WT-like)^1^** | **Effect estimate (95% CI)^2^** | **P** | **N (ref, WT-like)^1^** | **Effect estimate (95% CI)^2^** | **P** |
| Birth |  |  |  | 5336 (5315; 21) | -0.09 (-0.32, 0.14) | 0.45 |  |  |  |
| 4 months | 966 (962; 4) | 0.62 (-0.86, 2.09) | 0.41 | 1031 (1027; 4) | -0.02 (-0.78, 0.73) | 0.95 | 969 (965; 4) | -1.12 (-3.51, 1.27) | 0.36 |
| 8 months | 3297 (3281; 16) | -0.08 (-0.84, 0.68) | 0.84 | 3682 (3665; 17) | -0.20 (-0.67, 0.28) | 0.42 | 3365 (3348; 17) | -0.44 (-1.70, 0.81) | 0.49 |
| 12 months | 1366 (1359; 7) | -0.05 (-1.15, 1.05) | 0.93 | 1608 (1600; 8) | 0.17 (-0.62, 0.95) | 0.68 | 1382 (1375; 7) | 0.77 (-1.27, 2.82) | 0.46 |
| 18 months | 2713 (2698; 15) | 0.43 (-0.34, 1.20) | 0.28 | 2899 (2883; 16) | 0.001 (-0.61, 0.62) | 0.996 | 2791 (2776; 15) | -0.60 (-2.16, 0.97) | 0.45 |
| 2 years | 890 (886; 4) | 1.14 (-0.26, 2.55) | 0.11 | 924 (920; 4) | 0.75 (-0.66, 2.16) | 0.30 | 912 (908; 4) | -0.40 (-3.47, 2.67) | 0.80 |
| 2.5 years | 672 (669; 3) | 0.34 (-1.23, 1.91) | 0.67 | 697 (694; 3) | 0.37 (-1.49, 2.22) | 0.70 | 680 (677; 3) | 0.40 (-3.30, 4.09) | 0.83 |
| 3 years | 1051 (1048; 3) | 0.35 (-1.30, 2.00) | 0.68 | 1080 (1076; 4) | 0.09 (-1.66, 1.84) | 0.92 | 1096 (1093; 3) | -0.52 (-4.62, 3.58) | 0.80 |
| 3.5 years | 2434 (2423; 11) | -0.40 (-1.21, 0.40) | 0.33 | 2513 (2502; 11) | -0.33 (-1.44, 0.78) | 0.56 | 2517 (2506; 11) | 0.16 (-2.05, 2.38) | 0.89 |
| 4 years | 1109 (1105; 4) | 0.19 (-1.26, 1.64) | 0.80 | 1129 (1125; 4) | 0.06 (-2.07, 2.20) | 0.95 | 1152 (1148; 4) | -0.53 (-4.51, 3.45) | 0.79 |
| 5 years | 1281 (1276; 5) | -0.23 (-1.78, 1.32) | 0.77 | 1318 (1312; 6) | -0.75 (-3.03, 1.52) | 0.52 | 1380 (1375; 5) | -1.89 (-6.54, 2.76) | 0.43 |
| 8 years | 5226 (5208; 18) | -0.64 (-1.57, 0.29) | 0.17 | 5226 (5208; 18) | -1.98 (-4.11, 0.16) | 0.07 | 5231 (5213; 18) | -2.36 (-4.99, 0.28) | 0.08 |
| 9 years | 4437 (4419; 18) | -0.49 (-1.57, 0.59) | 0.37 | 4545 (4527; 18) | -1.55 (-4.19, 1.08) | 0.25 | 4699 (4681; 18) | -1.68 (-4.37, 1.01) | 0.22 |
| 10 years | 5360 (5339; 21) | -0.79 (-1.99, 0.41) | 0.20 | 5395 (5374; 21) | -2.16 (-5.29, 0.96) | 0.18 | 5363 (5342; 21) | -1.13 (-3.84, 1.57) | 0.41 |
| 11 years | 5032 (5014; 18) | -0.42 (-1.82, 0.98) | 0.56 | 5066 (5048; 18) | -1.94 (-5.74, 1.87) | 0.32 | 5041 (5023; 18) | -1.83 (-4.90, 1.24) | 0.24 |
| 12 years | 4913 (4895; 18) | -0.17 (-1.70, 1.37) | 0.83 | 4918 (4900; 18) | -1.23 (-5.75, 3.29) | 0.59 | 4915 (4897; 18) | -1.07 (-4.39, 2.26) | 0.53 |
| 13 years | 4635 (4618; 17) | -0.09 (-1.72, 1.53) | 0.91 | 4635 (4618; 17) | -0.98 (-6.04, 4.09) | 0.71 | 4680 (4663; 17) | -0.84 (-4.48, 2.81) | 0.65 |
| 14 years | 4339 (4321; 18) | 0.03 (-1.55, 1.61) | 0.97 | 4339 (4321; 18) | 0.20 (-4.99, 5.39) | 0.94 | 4344 (4326; 18) | 0.47 (-3.07, 4.02) | 0.79 |
| 15 years | 3852 (3837; 15) | -0.37 (-2.10, 1.36) | 0.68 | 3852 (3837; 15) | -0.89 (-6.63, 4.85) | 0.76 | 3858 (3843; 15) | 0.01 (-3.46, 3.49) | 0.99 |
| 18 years | 3489 (3478; 11) | -0.74 (-3.13, 1.66) | 0.55 | 3491 (3480; 11) | -2.10 (-9.65, 5.44) | 0.58 | 3492 (3481; 11) | -0.12 (-3.96, 3.72) | 0.95 |
| 24 years | 2691 (2681; 10) | -1.53 (-4.55, 1.48) | 0.32 | 2693 (2683; 10) | -5.80 (-15.19, 3.60) | 0.23 | 2693 (2683; 10) | -1.18 (-5.12, 2.77) | 0.56 |

*BMI = body mass index; CI = confidence interval; cLoF = complete loss of function; GoF = gain of function; LoF = loss of function; pLoF = partial loss of function; SD = standard deviation; WT = wild-type. ^1^N represents the total sample size in each analysis with numbers in brackets representing the number of individuals in the reference group (i.e., non-LoF carriers with synonymous, common variations or no LoF mutation and individuals with GoF mutations) and individuals carrying WT-like mutations, respectively. ^2^Estimates represent the change in each anthropometric trait (units in table) in carriers vs. the reference group (e.g., non-LoF carriers with synonymous, common variations, no LoF mutation or GoF mutations). Analyses were linear regression models adjusted for sex.*

**Supplementary Table 6.** Summary of measurements included in and model fit for BMI (kg/m^2^) trajectories

|  | **Summary of Measurements** | | |  | **Model fit for BMI trajectories** | | |
| --- | --- | --- | --- | --- | --- | --- | --- |
|  | **Number of participants with at least one measure of BMI^1^** | **Total number of measures** | **Median (IQR) measures per participant** | **Mean (SD) predicted intercept and slopes^2^** | **Mean predicted BMI in kg/m^2^ (SD)^3^** | **Mean observed BMI in kg/m^2^ (SD)** | **Mean difference between observed and predicted BMI in kg/m^2^ (95% CI)^4^** |
| Overall | 5716 | 45622 | 9 (8 to 9) | - | - | - | - |
| 18 months | 646 | 646 | 1 (1 to 1) | 17.04 (1.23) | 17.03 (1.23) | 17.12 (1.32) | 0.09 (-0.85, 1.03) |
| 18 months – 3.5 years | 691 | 2875 | 5 (4 to 5) | -0.28 (0.28) | 16.95 (1.27) | 16.75 (1.37) | -0.01 (-0.91, 0.89) |
| 3.5 – 5 years | 652 | 1270 | 2 (2 to 3) | -0.20 (0.33) | 16.26 (1.38) | 16.27 (1.45) | 0.01 (-0.76, 0.78) |
| 5 – 8 years | 4081 | 4281 | 1 (1 to 1) | 0.61 (0.54) | 16.17 (1.77) | 16.13 (1.93) | -0.04 (-0.81, 0.72) |
| 8 – 15 years | 5695 | 32361 | 7 (6 to 7) | 0.67 (0.32) | 18.74 (3.25) | 18.77 (3.37) | 0.01 (-1.33, 1.34) |
| 15 – 18 years | 3841 | 4835 | 1 (1 to 2) | 0.52 (0.97) | 23.67 (5.63) | 22.39 (3.93) | -0.02 (-1.41, 1.37) |

*BMI = body mass index; CI = confidence interval; IQR = inter-quartile range; SD = standard deviation*

*^1^Individuals who had full data on BMI and were in the sequence set*

*^2^The data at 18 months relate to predictions from the multilevel model at exactly 1.5 years (i.e., the intercept).
^3^The data at 18 months relate to the BMI measurement carried out at mean age 1.53 years.
^4^Range within which 95% of the differences between observed BMI measurements and those predicted by the multi-level model lie.*

**Supplementary Table 7.** Summary of measurements included in and model fit for weight (kg) trajectories

|  | **Summary of Measurements** | | |  | **Model fit for weight trajectories** | | |
| --- | --- | --- | --- | --- | --- | --- | --- |
|  | **Number of participants with at least one measure of weight^1^** | **Total number of measures** | **Median (IQR) measures per participant** | **Mean (SD) predicted intercept and slopes^2^** | **Mean predicted weight in kg (SD)^3^** | **Mean observed weight in kg (SD)** | **Mean difference between observed and predicted weight in kg (SD)^4^** |
| Overall | 5716 | 53083 | 10 (9 to 10) | - | - | - | - |
| Birth | 5354 | 5354 | 1 (1 to 1) | 3.51 (0.18) | 3.51 (0.18) | 3.43 (0.54) | -0.08 (-0.94, 0.78) |
| Birth – 12 months | 717 | 6913 | 2 (2 to 3) | 6.90 (0.87) | 7.94 (1.92) | 8.41 (1.67) | 0.48 (-0.73, 1.68) |
| 12 months – 8 years | 4168 | 8792 | 7 (1 to 8) | 2.28 (0.63) | 19.64 (6.34) | 19.73 (6.52) | 0.08 (-1.58, 1.74) |
| 8 – 15 years | 5696 | 32541 | 7 (6 to 7) | 4.69 (1.25) | 42.42 (12.93) | 42.51 (13.38) | -0.02 (-4.40, 4.37) |
| 15 – 18 years | 3843 | 4837 | 1 (1 to 2) | 2.88 (2.84) | 72.58 (20.40) | 65.79 (13.24) | 0.04 (-4.83, 4.91) |

*CI = confidence interval; IQR = inter-quartile range; SD = standard deviation.*

*^1^Individuals who had full data on weight and were in the sequence set*

*^2^The data at birth relate to predictions from the multilevel model at birth (i.e., the intercept).
^3^The data at birth relate to the weight measurement carried out at birth.
^4^Range within which 95% of the differences between observed weight measurements and those predicted by the multi-level model lie.*

**Supplementary Table 8.** Summary of measurements included in and model fit for height (cm) trajectories

|  | **Summary of Measurements** | | |  | **Model fit for height trajectories** | | |
| --- | --- | --- | --- | --- | --- | --- | --- |
|  | **Number of participants with at least one measure of height^1^** | **Total number of measures** | **Median (IQR) measures per participant** | **Mean (SD) predicted intercept and slopes^2^** | **Mean predicted height in cm (SD)^3^** | **Mean observed height in cm (SD)** | **Mean difference between observed and predicted height in cm (SD)^4^** |
| Overall | 5716 | 45962 | 9 (8 to 9) | - | - | - | - |
| 18 months | 647 | 647 | 1 (1 to 1) | 82.77 (2.64) | 82.77 (2.64) | 81.69 (2.72) | -1.08 (-2.68, 0.52) |
| 18 months – 5 years | 692 | 4149 | 6 (6 to 7) | 7.72 (0.69) | 88.79 (11.11) | 94.48 (9.12) | 0.18 (-1.81, 2.16) |
| 5 – 15 years | 5704 | 36971 | 7 (6 to 8) | 5.71 (0.47) | 145.78 (14.90) | 145.94 (15.32) | 0.09 (-4.04, 4.23) |
| 15 – 18 years | 3844 | 4842 | 1 (1 to 2) | -4.69 (4.25) | 159.91 (26.51) | 171.26 (9.16) | -0.95 (-7.60, 5.70) |

*CI = confidence interval; IQR = inter-quartile range; SD = standard deviation.*

*^1^Individuals who had full data on height and were in the sequence set*

*^2^The data at 18 months relate to predictions from the multilevel model at exactly 1.5 years (i.e., the intercept).
^3^The data at 18 months relate to the height measurement carried out at age 1.53 years.
^4^Range within which 95% of the differences between observed height measurements and those predicted by the multi-level model lie.*

**Supplementary Table 9.** Association between *MC4R* LoF of cAMP accumulation with predicted weight trajectory between the ages at birth and 18 years using linear spline multi-level models.

| **Intercept and slopes** | **Mean weight trajectory (95% CI) in reference group^1^** | **Difference in the intercept and slopes between ages with *MC4R* LoF mutation^2^** | |
| --- | --- | --- | --- |
|  |  | **Estimate (95% CI)** | **P-value** |
| Birth (kg) | 3.44 (3.42, 3.47) | 0.18 (-0.10, 0.47) | 0.21 |
| Change between birth – 12 months (kg/year) | 6.50 (6.39, 6.61) | 1.14 (-0.81, 3.09) | 0.25 |
| Change between 12 months – 8 years (kg/year) | 2.39 (2.36, 2.42) | 0.84 (0.40, 1.28) | 1.63x10^-04^ |
| Change between 8 – 15 years (kg/year) | 4.72 (4.66, 4.77) | 1.33 (0.66, 1.99) | 9.62x10^-05^ |
| Change between 15 – 18 years (kg/year) | 1.11 (0.98, 1.24) | 0.43 (-1.37, 2.23) | 0.64 |

*cLOF = complete loss of function; CI = confidence interval; GoF = gain of function; LoF = loss of function; pLoF = partial loss of function; WT-like = wild-type like.*

*^1^The reference group included all individuals with synonymous, common variations or no LoF mutation and individuals with GoF or WT-like mutations – the “non-LoF carriers”.*

*^2^Estimates represent the change in the intercept (kg) and slopes (kg/year) of weight in carriers vs. non-carriers of MC4R LoF mutations (i.e., individuals with synonymous, common variations or no LoF mutations and individuals with WT-like and GoF mutations). Analyses were adjusted for sex.*

**Supplementary Table 10.** Association between *MC4R* LoF of cAMP accumulation with predicted BMI trajectory between the ages of 18 months and 18 years using linear spline multi-level models.

| **Intercept and slopes** | **Mean BMI trajectory (95% CI) in reference group^1^** | **Difference in the intercept and slopes between ages with *MC4R* LoF mutation^2^** | |
| --- | --- | --- | --- |
|  |  | **Estimate (95% CI)** | **P-value** |
| 18 months (kg/m^2^) | 16.84 (16.72, 16.96) | 0.81 (-1.39, 3.00) | 0.47 |
| Change between 18 months – 3.5 years (kg/m^2^/year) | -0.23 (-0.27, -0.20) | 0.13 (-0.46, 0.71) | 0.67 |
| Change between 3.5 – 5 years (kg/m^2^/year) | -0.20 (-0.27, -0.13) | 0.75 (-0.43, 1.94) | 0.21 |
| Change between 5 – 8 years (kg/m^2^/year) | 0.65 (0.63, 0.68) | 0.39 (0.03, 0.75) | 0.04 |
| Change between 8 – 15 years (kg/m^2^/year) | 0.71 (0.70, 0.73) | 0.12 (-0.08, 0.32) | 0.25 |
| Change between 15 – 18 years (kg/m^2^/year) | 0.28 (0.20, 0.36) | 0.57 (-0.61, 1.74) | 0.34 |

*BMI = body mass index; CI = confidence interval; cLOF = complete loss of function; GoF = gain of function; LoF = loss of function; pLOF = partial loss of function.*

*^1^The reference group included all individuals with synonymous, common variations or no LoF mutation and individuals with GoF or WT-like mutations – the “non-LoF carriers”.*

*^2^Estimates represent the change in the intercept (kg/m^2^) and slopes (kg/m^2^/year) of BMI in carriers vs. non-carriers of MC4R LoF mutations (i.e., individuals with synonymous, common variations or no LoF mutations and individuals with WT-like and GoF mutations). Analyses were adjusted for sex.*

**Supplementary Table 11.** Association between *MC4R* LoF of cAMP accumulation with predicted height trajectory between the ages of 18 months and 18 years using linear spline multi-level models.

| **Intercept and slopes** | **Mean height trajectory (95% CI) in reference group^1^** | **Difference in the intercept and slopes between ages with *MC4R* LoF mutation^2^** | |
| --- | --- | --- | --- |
|  |  | **Estimate (95% CI)** | **P-value** |
| 18 months (cm) | 81.90 (81.70, 82.11) | 2.11 (-1.50, 5.73) | 0.25 |
| Change between 18 months – 5 years (cm/year) | 7.79 (7.74, 7.84) | 0.48 (-0.29, 1.26) | 0.23 |
| Change between 5 – 15 years (cm/year) | 5.50 (5.48, 5.53) | 0.07 (-0.23, 0.38) | 0.64 |
| Change between 15 – 18 years (cm/year) | -8.14 (-8.34, -7.95) | -2.27 (-5.12, 0.57) | 0.12 |

*cLoF = complete loss of function; CI = confidence interval; GoF = gain of function; LoF = loss of function; pLoF = partial loss of function; WT-like = wild-type like.*

*^1^The reference group included all individuals with synonymous, common variations or no LoF mutation and individuals with GoF or WT-like mutations – the “non-LoF carriers”.*

*^2^Estimates represent the change in the intercept (cm) and slopes (cm/year) of height in carriers vs. non-carriers of MC4R LoF mutations (i.e., individuals with synonymous, common variations or no LoF mutations and individuals with WT-like and GoF mutations). Analyses were adjusted for sex.*

**Supplementary Table 12.** Age-specific associations between *MC4R* LoF of β-arrestin-2 coupling and BMI, weight and height.

| **Age** | **BMI (kg/m^2^)** | | | **Weight (kg)** | | | **Height (kg)** | | |
| --- | --- | --- | --- | --- | --- | --- | --- | --- | --- |
|  | **N (ref, pLoF, cLoF)^1^** | **Effect estimate (95% CI)^2^** | **P** | **N (ref, pLoF, cLoF)^1^** | **Effect estimate (95% CI)^2^** | **P** | **N (ref, pLoF, cLoF)^1^** | **Effect estimate (95% CI)^2^** | **P** |
| Birth |  |  |  | 5354 (5341; 8; 5) | 0.25 (-0.04, 0.54) | 0.09 |  |  |  |
| 4 months | 967 (965; 2; 0) | -0.70 (-2.78, 1.39) | 0.51 | 1034 (1031; 3; 0) | 0.26 (-0.61, 1.13) | 0.56 | 970 (968; 2; 0) | 0.87 (-2.50, 4.25) | 0.61 |
| 8 months | 3307 (3301; 6; 0) | -0.56 (-1.79, 0.68) | 0.38 | 3690 (3683; 6; 1) | 0.02 (-0.71, 0.76) | 0.95 | 3375 (3369; 6; 0) | 0.08 (-2.03, 2.20) | 0.94 |
| 12 months | 1370 (1368; 2; 0) | 1.11 (-0.94, 3.16) | 0.29 | 1613 (1609; 3; 1) | 1.77 (0.66, 2.88) | 0.002 | 1386 (1384; 2; 0) | 1.04 (-2.80, 4.88) | 0.60 |
| 18 months | 2719 (2713; 4; 2) | 1.16 (-0.06, 2.38) | 0.06 | 2905 (2899; 4; 2) | 1.36 (0.36, 2.37) | 0.01 | 2797 (2791; 4; 2) | 1.66 (-0.81, 4.13) | 0.19 |
| 2 years | 891 (889; 2; 0) | 0.73 (-1.26, 2.71) | 0.47 | 925 (923; 2; 0) | 1.20 (-0.79, 3.19) | 0.24 | 913 (911; 2; 0) | 2.15 (-2.20, 6.49) | 0.33 |
| 2.5 years | 673 (671; 2; 0) | -0.09 (-2.01, 1.82) | 0.92 | 698 (696; 2; 0) | 0.79 (-1.48, 3.06) | 0.50 | 681 (679; 2; 0) | 3.07 (-1.46, 7.59) | 0.18 |
| 3 years | 1052 (1050; 2; 0) | -0.09 (-2.10, 1.93) | 0.93 | 1081 (1079; 2; 0) | 0.61 (-1.86, 3.08) | 0.63 | 1097 (1095; 2; 0) | 2.37 (-2.66, 7.39) | 0.36 |
| 3.5 years | 2443 (2436; 5; 2) | 0.23 (-0.78, 1.24) | 0.65 | 2522 (2515; 5; 2) | 1.42 (0.03, 2.82) | 0.04 | 2527 (2518; 6; 3) | 4.44 (1.99, 6.90) | 3.92x10^-04^ |
| 4 years | 1111 (1109; 2; 0) | 0.03 (-2.00, 2.06) | 0.98 | 1131 (1129; 2; 0) | 0.82 (-2.19, 3.83) | 0.59 | 1154 (1152; 2; 0) | 2.44 (-3.19, 8.07) | 0.40 |
| 5 years | 1284 (1281; 2; 1) | 0.77 (-1.23, 2.77) | 0.45 | 1321 (1318; 2; 1) | 2.55 (-0.67, 5.77) | 0.12 | 1383 (1380; 2; 1) | 3.73 (-2.28, 9.73) | 0.22 |
| 8 years | 5243 (5232; 6; 5) | 3.42 (2.23, 4.61) | 1.86x10^-08^ | 5243 (5232; 6; 5) | 8.19 (5.45, 10.92) | 4.75x10^-09^ | 5248 (5237; 6; 5) | 5.36 (1.99, 8.72) | 0.002 |
| 9 years | 4452 (4441; 6; 5) | 4.27 (2.89, 5.66) | 1.37x10^-09^ | 4560 (4549; 6; 5) | 11.56 (8.18, 14.93) | 2.10x10^-11^ | 4715 (4704; 6; 5) | 6.77 (3.34, 10.21) | 1.13x10^-04^ |
| 10 years | 5378 (5366; 7; 5) | 4.54 (2.94, 6.13) | 2.42x10^-08^ | 5412 (5400; 7; 5) | 13.94 (9.81, 18.08) | 4.32x10^-11^ | 5381 (5369; 7; 5) | 7.85 (4.27, 11.43) | 1.76x10^-05^ |
| 11 years | 5050 (5038; 8; 4) | 3.66 (1.95, 5.38) | 2.87x10^-05^ | 5084 (5072; 8; 4) | 11.38 (6.71, 16.04) | 1.79x10^-06^ | 5059 (5047; 8; 4) | 5.98 (2.21, 9.74) | 0.002 |
| 12 years | 4929 (4918; 7; 4) | 4.60 (2.64, 6.56) | 4.41x10^-06^ | 4934 (4923; 7; 4) | 16.85 (11.07, 22.63) | 1.15x10^-08^ | 4931 (4920; 7; 4) | 9.12 (4.87, 13.37) | 2.64x10^-05^ |
| 13 years | 4650 (4640; 6; 4) | 5.78 (3.67, 7.90) | 9.13x10^-08^ | 4650 (4640; 6; 4) | 19.60 (13.00, 26.21) | 6.24x10^-09^ | 4695 (4685; 6; 4) | 7.58 (2.82, 12.33) | 0.002 |
| 14 years | 4352 (4343; 5; 4) | 4.51 (2.27, 6.75) | 7.97x10^-05^ | 4352 (4343; 6; 4) | 18.82 (11.49, 26.15) | 5.03x10^-07^ | 4357 (4348; 5; 4) | 9.04 (4.03, 14.05) | 4.10x10^-04^ |
| 15 years | 3866 (3856; 6; 4) | 4.99 (2.87, 7.11) | 3.95x10^-06^ | 3866 (3856; 6; 4) | 20.78 (13.75, 27.81) | 7.38x10^-09^ | 3872 (3862; 6; 4) | 7.60 (3.34, 11.86) | 4.69x10^-04^ |
| 18 years | 3499 (3492; 4; 3) | 5.23 (2.23, 8.23) | 6.43x10^-04^ | 3501 (3494; 4; 3) | 21.76 (12.31, 31.22) | 6.63x10^-06^ | 3502 (3495; 4; 3) | 6.46 (1.64, 11.27) | 0.01 |
| 24 years | 2695 (2693; 1; 1) | 0.94 (-5.79, 7.67) | 0.79 | 2697 (2695; 1; 1) | 11.92 (-9.05, 32.89) | 0.27 | 2697 (2695; 1; 1) | 9.42 (0.61, 18.22) | 0.04 |

*BMI = body mass index; CI = confidence interval; cLoF = complete loss of function; GoF = gain of function; LoF = loss of function; pLoF = partial loss of function; SD = standard deviation; WT = wild-type. ^1^N represents the total sample size in each analysis with numbers in brackets representing the number of individuals in the reference group (i.e., individuals with synonymous, common variations or no LoF mutation and individuals with GoF or WT-like mutations – the “non-LoF carriers”), and individuals with pLoF or cLoF mutations, respectively. ^2^Estimates represent the change in each anthropometric trait (units in table) in carriers vs. non-carriers of MC4R LoF mutations. Analyses were linear regression models adjusted for sex.*

**Supplementary Table 13.** Age-specific associations between *MC4R* LoF of β-arrestin-2 coupling and WHR, fat mass and lean mass

| **Age** | **Fat mass (kg)** | | | **Lean mass(kg)** | | | **WHR** | | |
| --- | --- | --- | --- | --- | --- | --- | --- | --- | --- |
|  | **N (ref, pLoF, cLoF)^1^** | **Effect estimate (95% CI)^2^** | **P** | **N (ref, pLoF, cLoF)^1^** | **Effect estimate (95% CI)^2^** | **P** | **N (ref, pLoF, cLoF)^1^** | **Effect estimate (95% CI)^2^** | **P** |
| 8 years |  |  |  |  |  |  | 5078 (5067; 6; 5) | -0.003 (-0.03, 0.02) | 0.79 |
| 10 years | 5109 (5097; 7; 5) | 9.47 (6.70, 12.23) | 2.12x10^-11^ | 5109 (5097; 7; 5) | 3.88 (2.15, 5.62) | 1.19x10^-05^ | 5339 (5327; 7; 5) | 0.03 (0.01, 0.06) | 0.02 |
| 12 years | 4874 (4863; 7; 4) | 10.65 (6.78, 14.52) | 7.27x10^-08^ | 4874 (4863; 7; 4) | 5.55 (2.98, 8.11) | 2.31x10^­-05^ | 4926 (4915; 7; 4) | 0.04 (0.01, 0.07) | 0.01 |
| 14 years | 4295 (4286; 5; 4) | 12.21 (7.28, 17.15) | 1.27x10^‑06^ | 4295 (4286; 5; 4) | 5.93 (2.09, 9.77) | 0.002 |  |  |  |
| 15 years | 3750 (3740; 6; 4) | 14.42 (9.36, 19.47) | 2.40x10^-08^ | 3750 (3740; 6; 4) | 5.17 (1.75, 8.59) | 0.003 |  |  |  |
| 18 years | 3408 (3401; 4; 3) | 17.73 (10.69, 24.78) | 8.28x10^‑07^ | 3408 (3401; 4; 3) | 2.71 (-1.22, 6.65) | 0.18 |  |  |  |
| 24 years | 2631 (2629; 1; 1) | 11.90 (-2.49, 26.29) | 0.11 | 2631 (2629; 1; 1) | -0.45 (-9.21, 8.32) | 0.92 | 2692 (2690; 1; 1) | 0.04 (-0.04, 0.12) | 0.35 |

*CI = confidence interval; cLoF = complete loss of function; GoF = gain of function; LoF = loss of function; pLoF = partial loss of function; SD = standard deviation; WHR = waist-hip ratio WT = wild-type*

*^1^N represents the total sample size in each analysis with numbers in brackets representing the number of individuals in the reference group (i.e., individuals with synonymous, common variations or no LoF mutations and individuals with GoF or WT-like mutations – the “non-LoF carriers”), and individuals with pLoF or cLoF mutations, respectively.*

*^2^Estimates represent the change in each anthropometric trait (units in table) in carriers vs. non-carriers of MC4R LoF mutations. Analyses were linear regression models adjusted for sex.*

**Supplementary Table 14.** Age-specific associations between the weighted genome-wide polygenic risk score and BMI

| **Age of BMI measurement** | **N** | **Effect estimate (95% CI)^1^** | **P-value** |
| --- | --- | --- | --- |
| 4 months | 903 | 0.30 (-0.02, 0.63) | 0.06 |
| 8 months | 3012 | 0.13 (-0.06, 0.31) | 0.18 |
| 12 months | 1263 | 0.17 (-0.09, 0.44) | 0.20 |
| 18 months | 2478 | 0.11 (-0.10, 0.31) | 0.30 |
| 2 years | 810 | 0.18 (-0.14, 0.51) | 0.27 |
| 2.5 years | 629 | -0.02 (-0.35, 0.31) | 0.91 |
| 3 years | 968 | 0.28 (-0.02, 0.57) | 0.07 |
| 3.5 years | 2229 | 0.39 (0.20, 0.58) | 6.79x10^-05^ |
| 4 years | 1010 | 0.38 (0.09, 0.68) | 0.01 |
| 5 years | 1182 | 0.59 (0.27, 0.91) | 3.54x10^-05^ |
| 8 years | 4752 | 1.06 (0.87, 1.25) | 5.34x10^-27^ |
| 9 years | 4050 | 1.22 (0.98, 1.46) | 5.90x10^-23^ |
| 10 years | 4868 | 1.64 (1.38, 1.90) | 5.96x10^-34^ |
| 11 years | 4579 | 1.81 (1.52, 2.10) | 1.76x10^-33^ |
| 12 years | 4474 | 1.89 (1.56, 2.22) | 2.94x10^-29^ |
| 13 years | 4208 | 1.99 (1.64, 2.34) | 2.13x10^-28^ |
| 14 years | 3950 | 1.80 (1.43, 2.16) | 3.71x10^-22^ |
| 15 years | 3510 | 2.05 (1.67, 2.43) | 3.17x10^-25^ |
| 18 years | 3164 | 2.64 (2.17, 3.10) | 3.28x10^-28^ |
| 24 years | 2442 | 3.10 (2.46, 3.73) | 2.70x10^-21^ |

*BMI = body mass index; CI = confidence interval*

*^1^Estimates represent the change in BMI (kg/m^2^) between the top 10^th^ percentile vs. lower 90^th^ percentile of the weighted genome-wide polygenic risk score, restricted to only those in the sequence set. Analyses were linear regression models adjusted for sex.*

**Supplementary Table 15.** Summary of measurements included in and model fit for BMI (kg/m^2^) trajectories

|  | **Summary of Measurements** | | |  | **Model fit for BMI trajectories** | | |
| --- | --- | --- | --- | --- | --- | --- | --- |
|  | **Number of participants with at least one measure of BMI^1^** | **Total number of measures** | **Median (IQR) measures per participant** | **Mean (SD) predicted intercept and slopes^2^** | **Mean predicted BMI in kg/m^2^ (SD)^3^** | **Mean observed BMI in kg/m^2^ (SD)** | **Mean difference between observed and predicted BMI in kg/m^2^ (95% CI)^4^** |
| Overall | 5162 | 41445 | 9 (8 to 9) | - | - | - | - |
| 18 months | 596 | 596 | 1 (1 to 1) | 17.08 (1.24) | 17.08 (1.24) | 17.15 (1.33) | 0.07 (-0.70, 0.83) |
| 18 months – 3.5 years | 638 | 2667 | 5 (4 to 5) | -0.31 (0.38) | 16.97 (1.31) | 16.78 (1.38) | -0.01 (-0.82, 0.80) |
| 3.5 – 5 years | 602 | 1167 | 2 (2 to 3) | -0.34 (0.22) | 16.24 (1.39) | 16.30 (1.48) | 0.06 (-0.67, 0.78) |
| 5 – 8 years | 3557 | 3735 | 1 (1 to 1) | 0.28 (0.55) | 16.24 (1.92) | 16.13 (1.92) | -0.11 (-1.06, 0.84) |
| 8 – 15 years | 5144 | 29498 | 7 (6 to 7) | 0.67 (0.31) | 18.70 (3.23) | 18.74 (3.36) | 0.01 (-1.44, 1.46) |
| 15 – 18 years | 3475 | 4378 | 1 (1 to 2) | 0.58 (0.49) | 23.83 (4.62) | 22.39 (3.90) | -0.01 (-1.51, 1.49) |

*BMI = body mass index; CI = confidence interval; IQR = inter-quartile range; SD = standard deviation.*

*^1^Individuals who had full data on BMI and were in the sequence set*

*^2^The data at 18 months relate to predictions from the multilevel model at exactly 1.5 years (i.e., the intercept).
^3^The data at 18 months relate to the BMI measurement carried out at a mean age of 1.53 years.*

**Supplementary Table 16.** Association between the weighted genome-wide polygenic risk score (lower 90^th^ and upper 10^th^ percentiles) with predicted BMI trajectory between the ages of 18 months and 18 years in the sequence set using linear spline multi-level models.

| **Intercept and slopes** | **Mean BMI trajectory (95% CI) in reference group^1^** | **Difference in the intercept and slopes between ages with the genome-wide polygenic risk score (upper 10^th^ vs. lower 90^th^ percentile)^2^** | |
| --- | --- | --- | --- |
|  |  | **Estimate (95% CI)** | **P-value** |
| 18 months (kg/m^2^) | 16.63 (16.49, 16.77) | -0.09 (-0.39, 0.21) | 0.55 |
| Change between 18 – 3.5 years (kg/m^2^/year) | -0.26 (-0.31, -0.20) | 0.10 (-0.02, 0.22) | 0.10 |
| Change between 3.5 – 5 years (kg/m^2^/year) | -0.34 (-0.40, -0.27) | 0.19 (0.06, 0.32) | 0.01 |
| Change between 5 – 8 years (kg/m^2^/year) | 0.28 (0.23, 0.33) | 0.31 (0.21, 0.41) | 3.22x10^-09^ |
| Change between 8 – 15 years (kg/m^2^/year) | 0.71 (0.69, 0.72) | 0.14 (0.10, 0.18) | 2.74x10^-14^ |
| Change between 15 – 18 years (kg/m^2^/year) | 0.49 (0.45, 0.53) | 0.10 (0.003, 0.20) | 0.04 |

*BMI = body mass index; CI = confidence interval*

*^1^The reference group included individuals in the lower 90^th^ percentile of the weighted genome-wide polygenic risk score*

*^2^Adjusted for sex*

**Supplementary Table 17.** Age-specific associations between *MC4R* LoF of cAMP accumulation and BMI, adjusting for the weighted genome-wide polygenic risk score

| **Age** | **BMI (kg/m^2^)** | | |
| --- | --- | --- | --- |
|  | **N^1^** | **Effect estimate (95% CI)^2^** | **P** |
| Birth |  |  |  |
| 4 months | 903 | -0.25 (-3.22, 2.72) | 0.87 |
| 8 months | 3010 | 0.08 (-1.06, 1.23) | 0.89 |
| 12 months | 1261 | 1.59 (-0.46, 3.64) | 0.13 |
| 18 months | 2477 | 0.85 (-0.63, 2.33) | 0.26 |
| 2 years | 810 | -0.32 (-3.13, 2.48) | 0.82 |
| 2.5 years | 629 | -0.63 (-3.34, 2.07) | 0.65 |
| 3 years | 969 | -0.54 (-3.39, 2.30) | 0.71 |
| 3.5 years | 2228 | 0.07 (-1.02, 1.15) | 0.91 |
| 4 years | 1010 | -0.61 (-2.61, 1.39) | 0.55 |
| 5 years | 1181 | 0.35 (-2.06, 2.76) | 0.78 |
| 8 years | 4752 | 1.79 (0.65, 2.94) | 0.002 |
| 9 years | 4050 | 2.58 (1.19, 3.97) | 2.76x10^-04^ |
| 10 years | 4868 | 2.77 (1.25, 4.28) | 3.40x10^-04^ |
| 11 years | 4579 | 2.34 (0.78, 3.90) | 0.003 |
| 12 years | 4474 | 2.77 (0.91, 4.64) | 0.004 |
| 13 years | 4208 | 3.77 (1.75, 5.79) | 2.50x10^-04^ |
| 14 years | 3950 | 2.21 (0.08, 4.35) | 0.04 |
| 15 years | 3510 | 3.74 (1.72, 5.77) | 3.01x10^-04^ |
| 18 years | 3164 | 4.46 (1.64, 7.28) | 0.002 |
| 24 years | 2442 | -0.34 (-6.70, 6.01) | 0.92 |

*BMI = body mass index; CI = confidence interval; cLoF = complete loss of function; GoF = gain of function; LoF = loss of function; pLoF = partial loss of function; SD = standard deviation; WT = wild-type.*

*^1^N represents the total sample size in each analysis.*

*^2^Estimates represent the change in each anthropometric trait (units in table) in carriers (i.e., individuals with pLoF or cLoF mutations) vs. non-carriers (i.e., individuals with synonymous, common variations or no LoF mutation, and individuals with GoF or WT-like mutations) of MC4R LoF mutations, after adjusting for sex and the weighted genome-wide polygenic risk score.*

**Supplementary Table 18.** Primers used for Human *MC4R* Sequencing

| **Primer name** | **Sequence (5'-3')** | **Chromosome 18 position^1^** | | **Strand** |
| --- | --- | --- | --- | --- |
| MC4R Exon_Forward | GGGGGACACTGGAATTCTCC | 60372357 | 60372376 | Negative |
| MC4R Exon_Reverse | ACCCTACACGGAAGAGAAAGC | 60371247 | 60371267 | Positive |
| MC4R_Sequencing_1_Forward | ACTGGAATTCTCCTGCCAGC | 60372350 | 60372369 | Negative |
| MC4R_Sequencing_1_Reverse | CCAACAAGCTGATGACACCC | 60372169 | 60372188 | Positive |
| MC4R_Sequencing_2_Forward | TGACTCTGGGTGTCATCAGC | 60372176 | 60372195 | Negative |
| MC4R_Sequencing_2_Reverse | TGGATGCAAGCAAGGAGCTA | 60371941 | 60371960 | Positive |
| MC4R_Sequencing_3_Forward | ACTCGGTGATCTGTAGCTCCT | 60371953 | 60371973 | Negative |
| MC4R_Sequencing_3_Reverse | GAAGCCATGAGAGCCAGCAT | 60371721 | 60371740 | Positive |
| MC4R_Sequencing_4_Forward | TGTTCTTCACCATGCTGGCT | 60371732 | 60371751 | Negative |
| MC4R_Sequencing_4_Reverse | GTGAGACATGAAGCACACACA | 60371501 | 60371521 | Negative |
| MC4R_Sequencing_5_Reverse | CACGGAAGAGAAAGCTGTTGC | 60371253 | 60371273 | Positive |

*^1^Assembly: GRCh38p13*

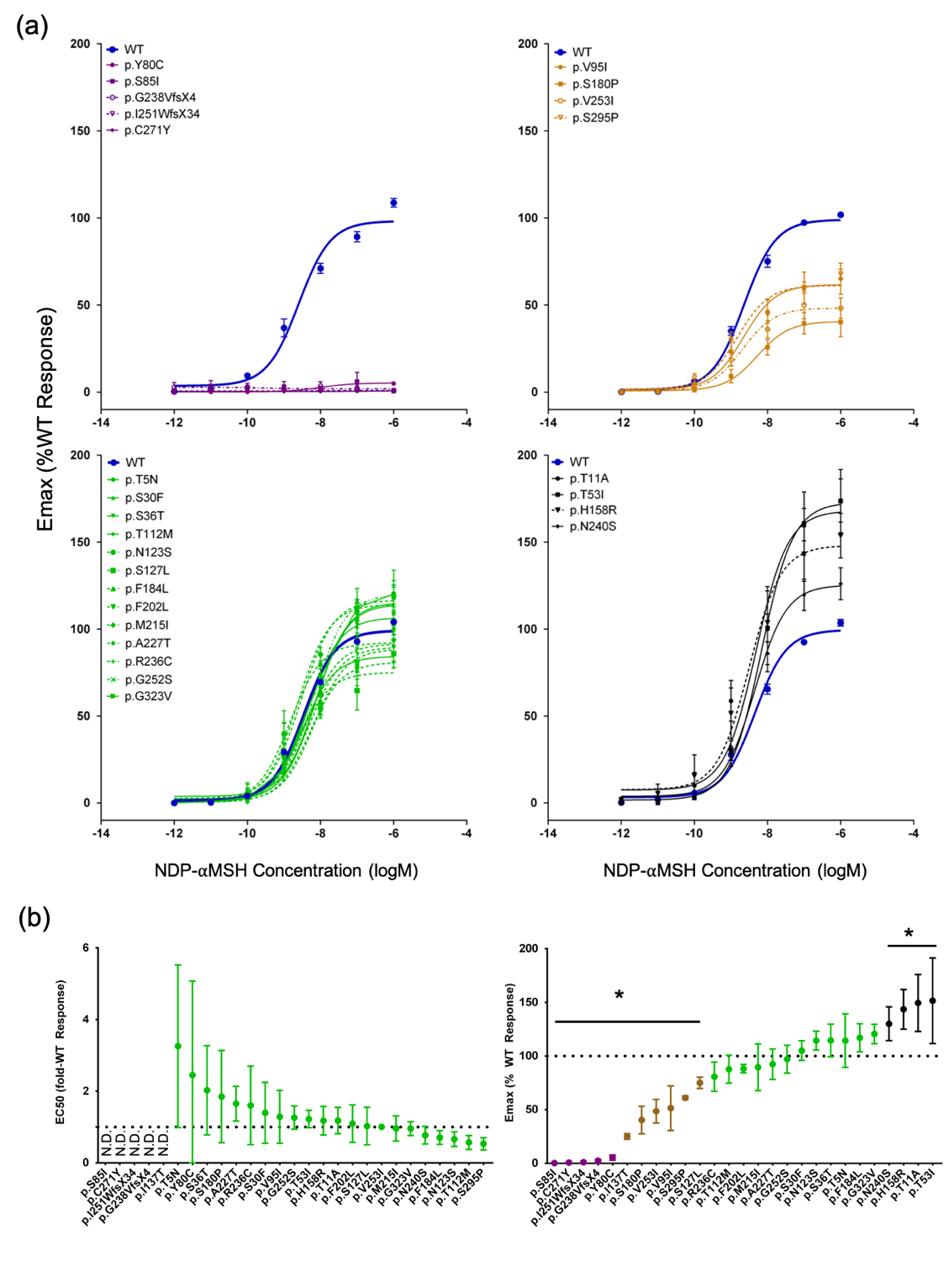
**Supplementary Figure 1.** β-arrestin-2 functional classification of *MC4R* variants

*cLoF = complete loss of function; GoF = gain of function; N.D. = not determined; NDP-αMSH = [Nle4,D-Phe7]-α-melanocyte-stimulating hormone; pLoF = partial loss of function; WT-like = wild-type like.*

*Colours represent cLoF (purple), pLoF (light brown), WT-like (green) and GoF (black). β-arrestin-2 coupling activity of MC4R mutations were characterised using a protein-protein interaction assay. (a) Dose response curves of MC4R mutants upon activation by NDP-MSH grouped by cLoF, pLoF, WT-like and GoF compared wild-type MC4R. (b) The relative EC_50_ (fold WT response, left panel) and E_max_ (% WT response right panel) were determined for each mutants and are presented in mean and 95% CI. * indicates p<0.05 by paired t-test.*

**Supplementary Figure 2.** Mean weight across time with *MC4R* LoF of cAMP accumulation

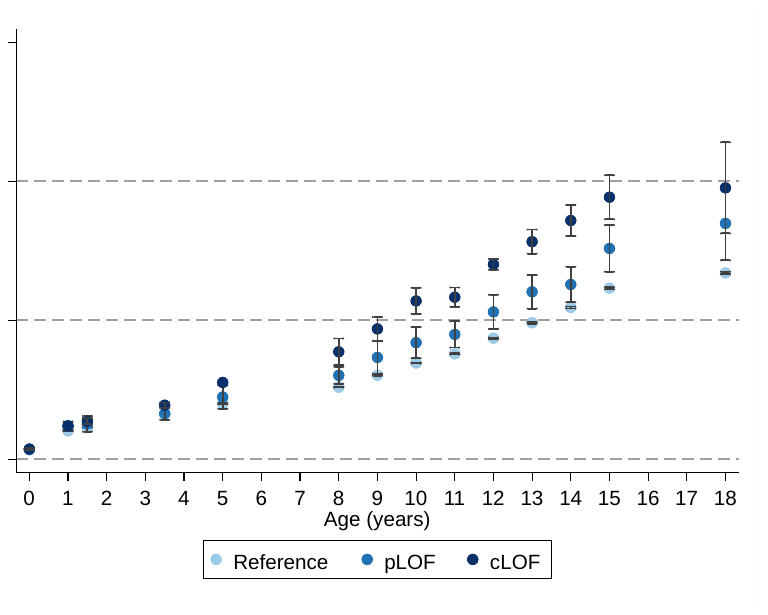

*cLoF = complete loss of function; GoF = gain of function; LoF = loss of function; pLoF = partial loss of function; WT = wild-type.*

*Figure shows mean weight at different age with MC4R LoF of cAMP (carriers of pLoF and cLoF) and the reference group (i.e., non-LoF carriers – combining individuals with synonymous, common variations or no LoF mutations and individuals with WT-like and GoF mutations). Figures only show results where all mutational groups (i.e., WT-like, GoF, pLoF and cLoF mutations) were represented by at least one individual at all time points between birth and 24 years.*

**Supplementary Figure 3.** Mean height across time with *MC4R* LoF of cAMP accumulation

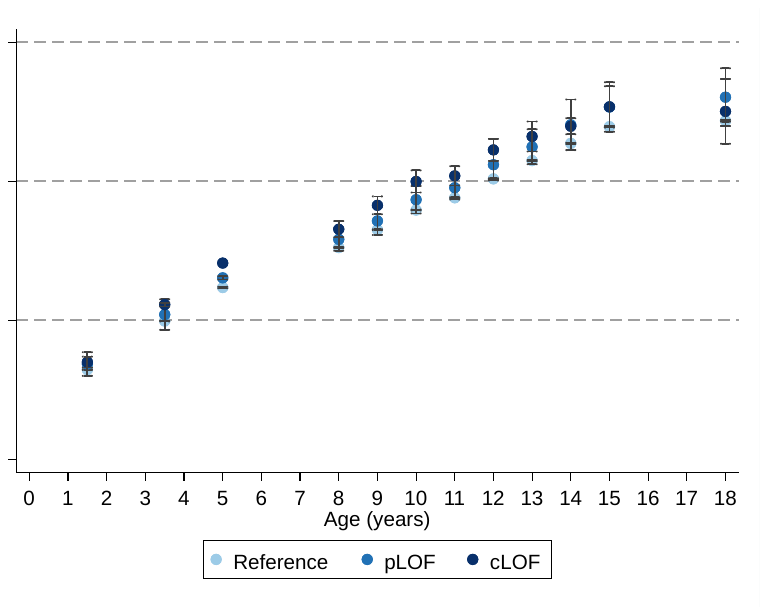

*cLoF = complete loss of function; GoF = gain of function; LoF = loss of function; pLoF = partial loss of function; WT = wild-type.*

*Figure shows mean height at different age with MC4R LoF of cAMP (carriers of pLoF and cLoF) and the reference group (i.e., non-LoF carriers – combining individuals with synonymous, common variations or no LoF mutations and individuals with WT-like and GoF mutations). Figures only show results where all mutational groups (i.e., WT-like, GoF, pLoF and cLoF mutations) were represented by at least one individual at all time points between birth and 24 years.*

**Supplementary Figure 4a.** Association between *MC4R* LoF of cAMP accumulation and arterial SBP across age in a model adjusting only for sex and a model adjusted for sex and BMI at the same age.

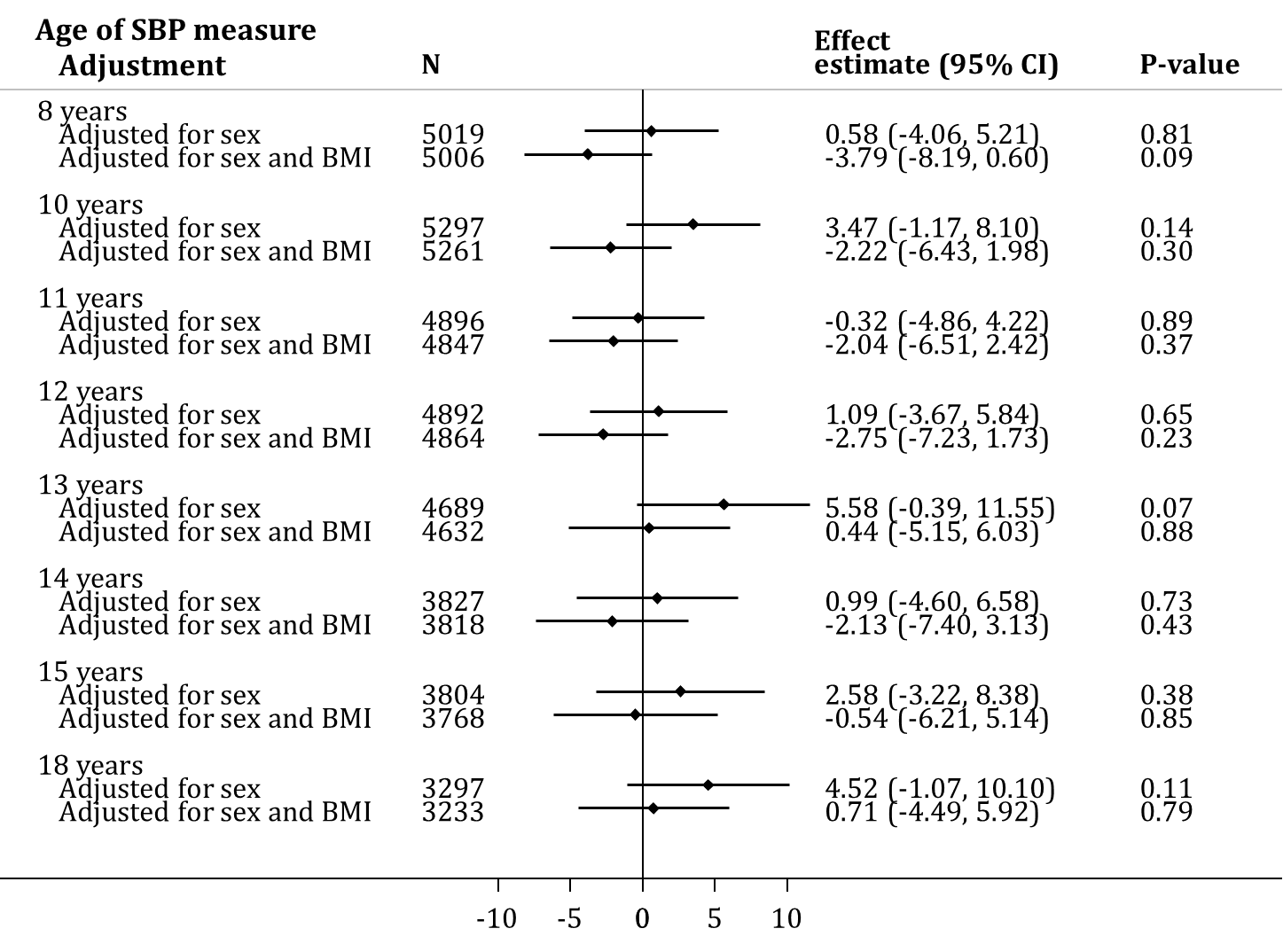

*BMI = body mass index; CI = confidence interval; cLoF = complete loss of function; GoF = gain of function; LoF = loss of function; pLoF = partial loss of function; SBP = systolic blood pressure; WT-like = wild-type like.*

*Estimates represent the change in SBP (mmHg) in carriers (i.e., pLoF or cLoF) vs. non-LoF carriers (i.e., individuals with synonymous, common variations or no LoF mutations and individuals with WT-like and GoF mutations) of MC4R LoF mutations.*

**Supplementary Figure 4b.** Association between *MC4R* LoF of cAMP accumulation and arterial DBP across age in a model adjusting only for sex and a model adjusted for sex and BMI at the same age.

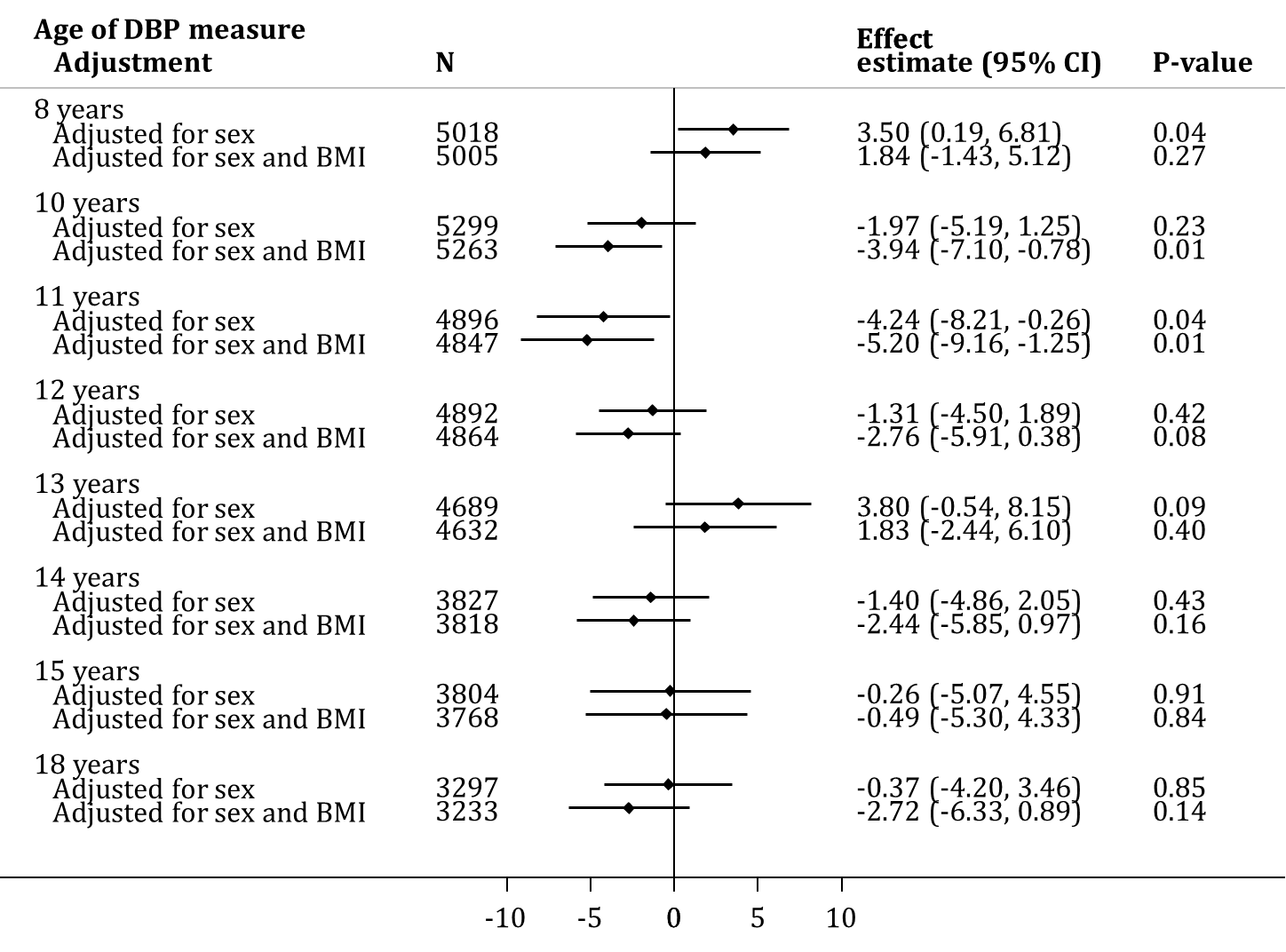

*BMI = body mass index; CI = confidence interval; cLoF = complete loss of function; DBP = diastolic blood pressure; GoF = gain of function; LoF = loss of function; pLoF = partial loss of function; WT-like = wild-type like.*

*Estimates represent the change in DBP (mmHg) in carriers (i.e., pLoF or cLoF) vs. non-LoF carriers (i.e, individuals with synonymous, common variations or no LoF mutations and individuals with WT-like and GoF mutations) of MC4R LoF mutations.*

**Supplementary Figure 5.** Association of *MC4R* LoF of cAMP accumulation with both central BP and LVMI at age 18 years in a model adjusting only for sex and a model adjusted for sex and BMI at the same age.

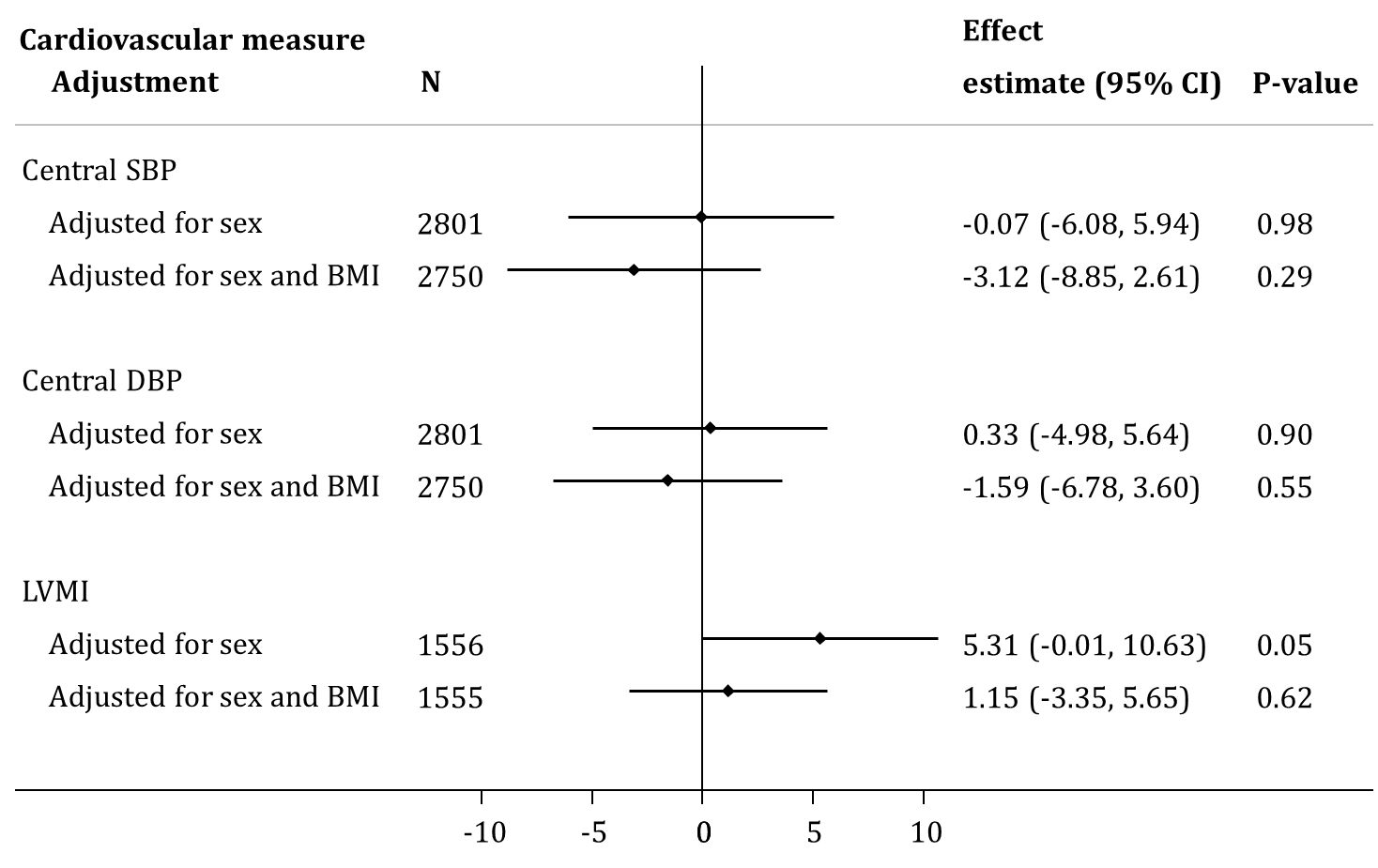

*BMI = body mass index; CI = confidence interval; cLoF = complete loss of function; DEBP = diastolic blood pressure; GoF = gain of function; LoF = loss of function; LVMI = left ventricular mass index; pLoF = partial loss of function; SBP = systolic blood pressure; WT-like = wild-type like.*

*Estimates represent the change in central DBP (mmHg), central SBP (mmHg) and LVMI (g/m^2.7^) in carriers (i.e., pLoF or cLoF) vs. non-LoF carriers (i.e., individuals with synonymous, common variations or no LoF mutations and individuals with WT-like and GoF mutations) of MC4R LoF mutations.*

**Supplementary Figure 6.** Association between *MC4R* LoF of cAMP accumulation with weight trajectory between birth and 18 years using linear spline multi-level models.

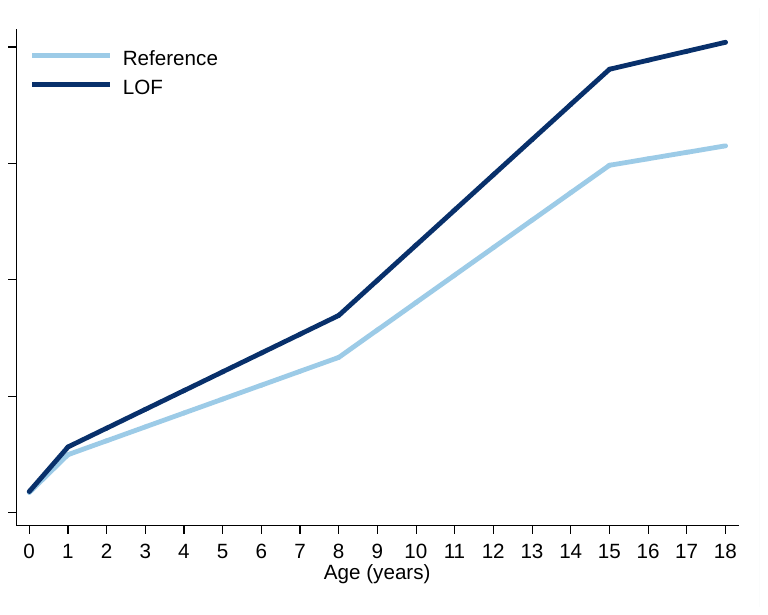

*cLoF = complete loss of function; GoF = gain of function; LoF = loss of function; pLoF = partial loss of function; WT-like = wild-type like.*

*Values for the reference group (i.e., all individuals with synonymous, common variations or no LoF mutations and individuals with WT-like and GoF mutations) and LoF mutations (i.e., combining pLoF and cLoF mutations) are depicted in light and dark blue, respectively. Effect estimates and confidence intervals of these analyses are presented in Supplementary Table 9.*

**Supplementary Figure 7.** Association between *MC4R* LoF of cAMP accumulation with height trajectory between the ages of 18 months and 18 years using linear spline multi-level models.

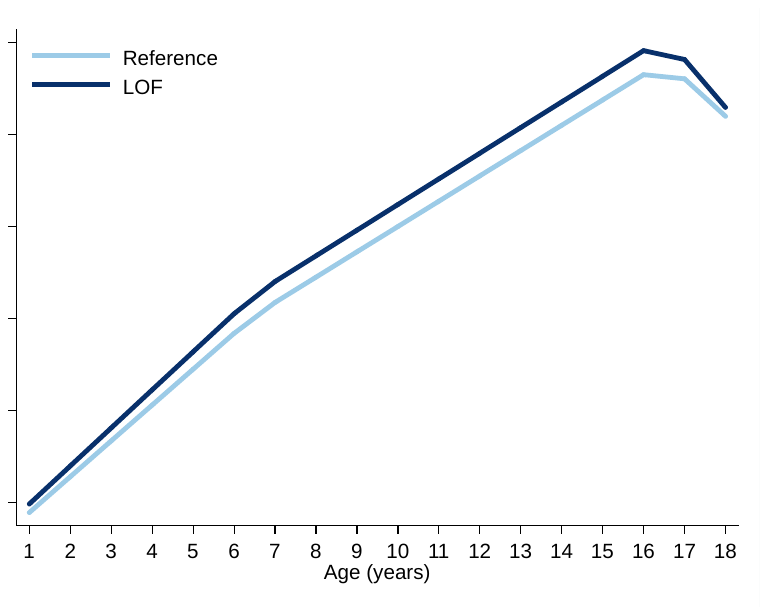
*cLOF = complete loss of function; GoF = gain of function; LoF = loss of function; pLoF = partial loss of function; WT-like = wild-type like.*

*Values for the reference group (i.e., all individuals with synonymous, common variations or no LoF mutations and individuals with WT-like and GoF mutations) and LoF mutations (i.e., combining pLoF and cLoF mutations) are depicted in light and dark blue, respectively. Effect estimates and confidence intervals of these analyses are presented in Supplementary Table 11.*

**Supplementary Figure 8.** Mean BMI across time with *MC4R* LoF of β-arrestin-2 coupling

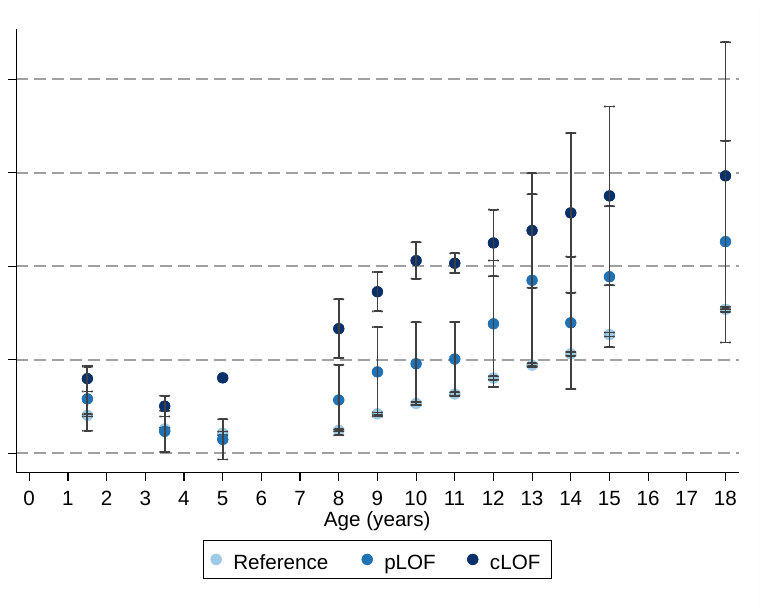

*cLoF = complete loss of function; GoF = gain of function; LoF = loss of function; pLoF = partial loss of function; WT = wild-type.*

*Figure shows mean BMI at different age with MC4R LoF of β-arrestin-2 (carriers of pLoF and cLoF) and the reference group (i.e., non-LoF carriers – combining individuals with synonymous, common variations or no LoF mutations and individuals with WT-like and GoF mutations). Figures only show results where all mutational groups (i.e., WT-like, GoF, pLoF and cLoF mutations) were represented by at least one individual at all time points between birth and 24 years.*

**Supplementary Figure 9.** Mean weight across time with *MC4R* LoF of β-arrestin-2 coupling

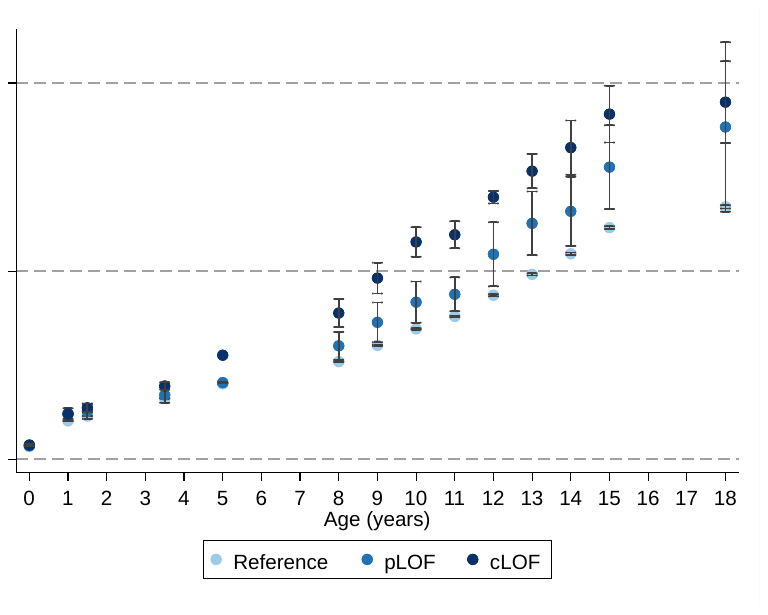

*cLoF = complete loss of function; GoF = gain of function; LoF = loss of function; pLoF = partial loss of function; WT = wild-type.*

*Figure shows mean weight at different age with MC4R LoF of β-arrestin-2 (carriers of pLoF and cLoF) and the reference group (i.e., non-LoF carriers – combining individuals with synonymous, common variations or no LoF mutations and individuals with WT-like and GoF mutations). Figures only show results where all mutational groups (i.e., WT-like, GoF, pLoF and cLoF mutations) were represented by at least one individual at all time points between birth and 24 years.*

**Supplementary Figure 10.** Mean height across time with *MC4R* LoF of β-arrestin-2 coupling

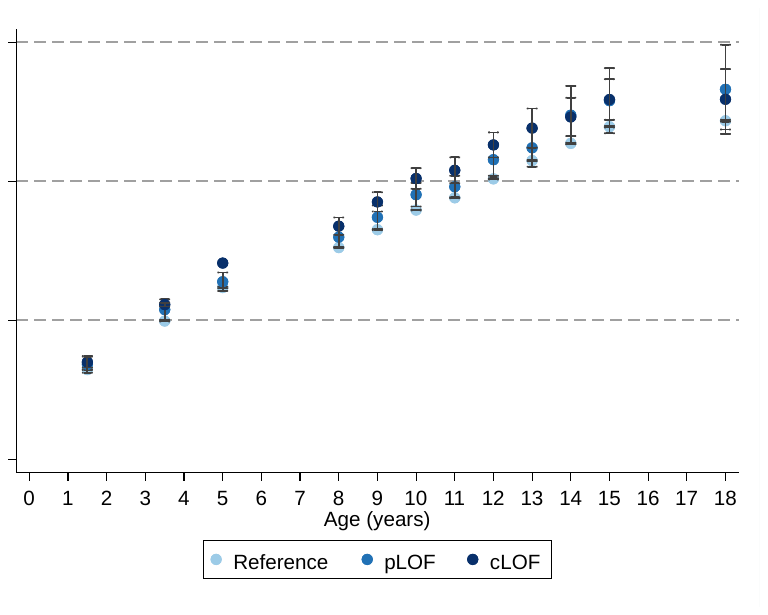

*cLoF = complete loss of function; GoF = gain of function; LoF = loss of function; pLoF = partial loss of function; WT = wild-type.*

*Figure shows mean height at different age with MC4R LoF of β-arrestin-2 (carriers of pLoF and cLoF) and the reference group (i.e., non-LoF carriers – combining individuals with synonymous, common variations or no LoF mutations and individuals with WT-like and GoF mutations). Figures only show results where all mutational groups (i.e., WT-like, GoF, pLoF and cLoF mutations) were represented by at least one individual at all time points between birth and 24 years.*

**Supplementary Figure 11.** Next generation sequencing of *MC4R*

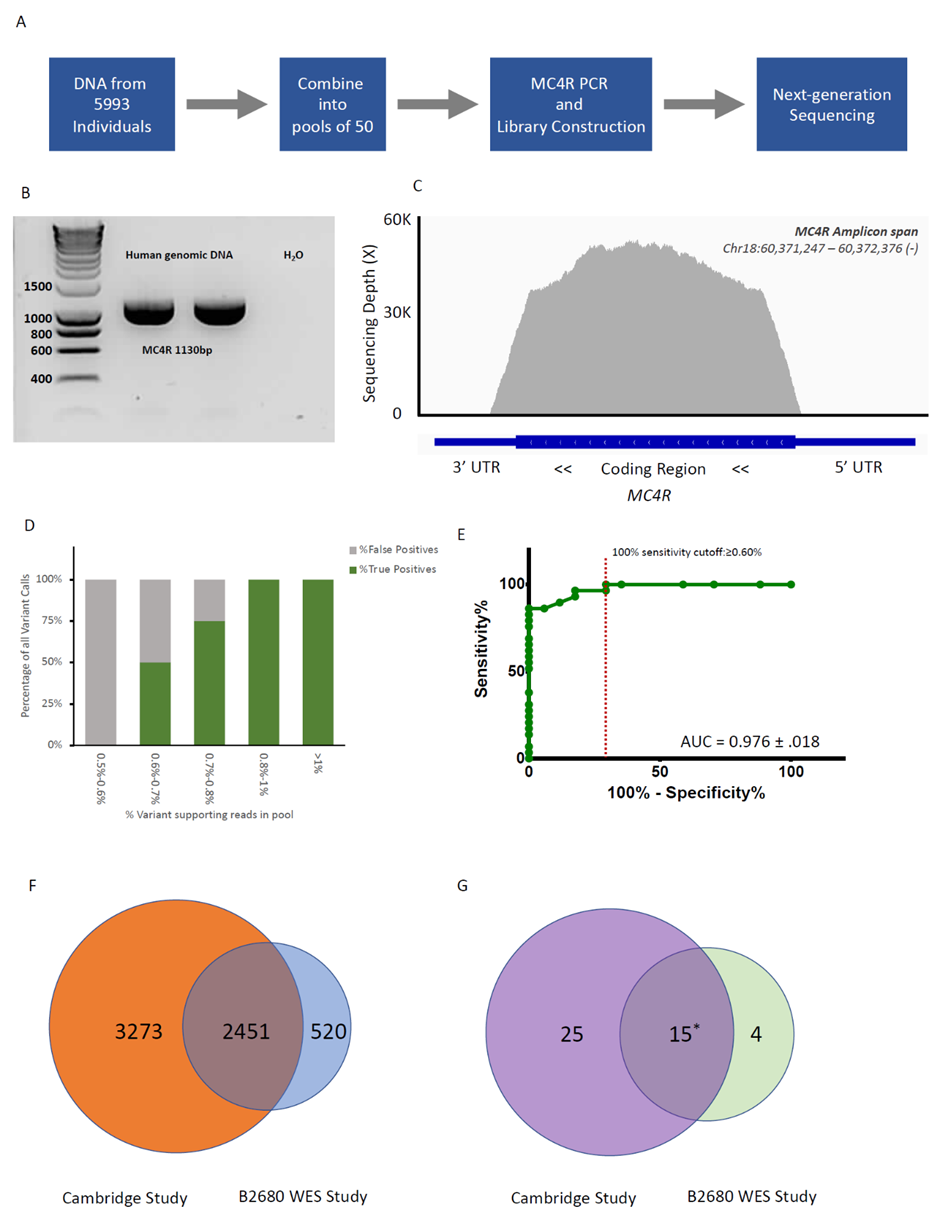

*ALSPAC = Avon Longitudinal Study of Parents and Children; AUC = area under the curve; NGS = next generation sequencing; WES = whole-exome sequencing.*

*(a) Pooled MC4R exon sequencing workflow for DNA samples from the ALSPAC cohort; (b) Agarose gel showing a 1130bp PCR product using MC4R exon primers (Supplementary Table 1); (c) A plot showing the sequencing coverage of MC4R coding region from a representative pool. The average per-base sequencing depth for all pools was 43,654 ± 356-fold; (d) The percentage of true positive and false positive calls binned by variant allele frequency (VAF) detected in pools; (e) Receiver operating curve analysis of VAF and call accuracy; (f) Comparison of participants included in the current study and another whole-exome sequencing (WES) study (with ALSPAC project number: B2680); (g) Comparison of non-synonymous variant carriages between this study and WES study. Of 40 mutational carriages found in Cambridge, 15 were found in both studies.  25 carriers found in the Cambridge study were not part of the WES study and 4 carriers found only in the WES study were not part of the Cambridge study.  *p.V103I and p.I251L were excluded in the analyses.*
